# Epidemic models characterize seizure propagation and the effects of epilepsy surgery in individualized brain networks based on MEG and invasive EEG recordings

**DOI:** 10.1101/2021.09.20.21263459

**Authors:** Ana. P. Millán, Elisabeth C.W. van Straaten, Cornelis J. Stam, Ida A. Nissen, Sander Idema, Johannes C. Baayen, Piet Van Mieghem, Arjan Hillebrand

## Abstract

**Background:** Epilepsy surgery is the treatment of choice for drug-resistant epilepsy patients. However, seizure-freedom is currently achieved in only 2*/*3 of the patients after surgery. In this study we have developed an individualized computational model based on functional brain networks to explore seizure propagation and the efficacy of different virtual resections. Eventually, the goal is to obtain individualized models to optimize resection strategy and outcome.

**Methods:** We have modelled seizure propagation as an epidemic process using the susceptible-infected (SI) model on individual functional networks derived from presurgical MEG. We included 10 patients who had received epilepsy surgery and for whom the surgery outcome at least one year after surgery was known. The model parameters were tuned in order to reproduce the patient-specific seizure propagation patterns as recorded with invasive EEG. We defined a personalized search algorithm that combined structural and dynamical information to find resections that maximally decreased seizure propagation for a given resection size. The optimal resection for each patient was defined as the smallest resection leading to at least a 90% reduction in seizure propagation.

**Results:** The individualized model reproduced the basic aspects of seizure propagation for 9 out of 10 patients when using the resection area as the origin of epidemic spreading, and for 10 out of 10 patients with an alternative definition of the seed region. We found that, for 7 patients, the optimal resection was smaller than the resection area, and for 4 patients we also found that a resection smaller than the resection area could lead to a 100% decrease in propagation. Moreover, for two cases these alternative resections included nodes outside the resection area.

**Conclusion:** Epidemic spreading models fitted with patient specific data can capture the fundamental aspects of clinically observed seizure propagation, and can be used to test virtual resections *in silico*. Combined with optimization algorithms, smaller or alternative resection strategies, that are individually targeted for each patient, can be determined with the ultimate goal to improve surgery outcome.

## 1 Introduction

Epilepsy is one of the most common neurological disorders, affecting between 4 and 10 per 1000 people worldwide [1]. There is not one single cause of epilepsy: it often occurs as an associated symptom of an underlying disease, but many other times it is produced by unknown causes [2]. This complicates the understanding of seizure dynamics, and to this day the microscopic mechanisms that lead to seizure generation and propagation are not fully understood [3]. It is generally assumed that a shift from normal neuronal activity to excessive synchronization [4] occurs due to decreased inhibition [5], but the actual nature of this transition is not clear.

Epilepsy is initially treated by anti-epileptic drugs (AEDs), but this approach is not effective for roughly 1 out of 3 people [6]. For these drug-resistant patients, epilepsy surgery is an optional treatment if a focal origin of the seizures can be found. The surgery then aims to remove or disconnect the brain regions thought to be involved in seizure generation. Currently, seizure freedom is achieved in up to 2 out of 3 patients who undergo epilepsy surgery, although surgery outcome varies greatly depending on epilepsy type [7]. Even when surgery is not completely successful, the majority of patients will still experience a reduction in seizure frequency or intensity after surgery. However, side-effects and cognitive complaints can also occur after surgery. These depend on the brain areas resected, but are difficult to predict accurately [8].

Traditionally, efforts to improve epilepsy surgery have aimed to better characterize the epileptogenic zone (EZ, defined as the minimal brain area or areas that need to be removed or disconnected to achieve seizure freedom [9]; by definition, this can only be confirmed after surgery, and prior to it only a hypothesis can be made). However, in recent years – and in line with the increasingly common view of the brain as a complex network – epilepsy is seen as a network disorder. Attention is thus shifting towards the definition of *epileptogenic networks* that can capture more details of seizure dynamics and the distribution of epileptiform activity [10–12]. An increasing number of findings support this perspective: topological properties of epileptogenic brain networks have been found to deviate from those seen in healthy controls [13, 14], and abnormal patterns of functional connectivity emerge [15–17] (although the results of different imaging modalities are sometimes contradictory [15–20]). A common finding in pathological brain networks is the association of disease with network hubs [21, 22]. In the case of epilepsy, hubs may facilitate the propagation of epileptiform activity to the rest of the brain [23–25]. Moreover, several studies have pointed out the existence of *pathological hubs*: abnormal, hyperconnected regions in the vicinity of the epileptic focus, which may facilitate seizure propagation [25–27].

Importantly, the network perspective of epilepsy implies that the effect of a resection cannot be predicted directly from the location of the removed region alone [28]: local resections can have widespread effects, but at the same time might not prevent the epileptogenic network from forming a new EZ eventually [29]. Computer models are then necessary to help predict the effect of a given resection [30]. Integrating patient specific data of different modalities, computer models allow us to test different resections *in silico* – i.e. using virtual resections – together with different markers of the EZ. Within this framework, Hebbink et al. [31] showed that the resection of the pathological network node is not necessarily the best approach to alleviate seizures, whereas Lopes et al. [32] found that the fraction of resected rich-club nodes correlated with surgery outcome. Going beyond topological network analysis, the simulation of ictal activity on top of brain networks can aid the identification of the EZ and prediction of surgery outcome, as well as predict possible side-effects. Such computational models can be used to identify epileptogenic areas [33, 34] or analyze different resection strategies [26, 32, 35–38], such that patient-specific resection strategies, that may lead to a better outcome or fewer side-effects than the standard surgery, can be tested [33, 39–41]. Validation of the models is usually attempted by looking for differences in the model predictions between seizure free and non-seizure free patients [42, 43] or by correlating seizure propagation on the model with the empirical data [44].

A basic consideration in the model definition is thus the nature of the underlying network. The temporal and spatial resolution of the resulting network-based model, and the interpretation of the connections between regions, will depend on the modality that was used to define the network structure. Studies on epileptogenic networks have considered both functional [32, 35, 37, 42, 43, 45] and structural [33, 36, 40, 41] networks, as they are both affected in patients with epilepsy [40–43]. However, functional networks can capture abnormalities in brain activity even in the absence of structural abnormalities [46]. Functional networks based on intracranial recordings [32, 35, 37, 42, 43] usually include ictal data and allow for highly precise characterization of some brain areas, however spatial sampling is sparse and biased due to an a priori hypothesis of the EZ, which may lead to bias in the analysis. Moreover, these invasive recordings are not always part of the presurgical evaluation. Non-invasive methods, such as Electro- and Magneto-Encephalography (EEG/MEG) have no risks of complications [47]. MEG is less affected by the skull and other tissue in the head, is reference-free and has higher spatial resolution than clinical scalp EEG [48]. This allows for a more accurate estimation of functional interactions between brain regions, and thus a more accurate reconstruction of the functional networks. MEG interictal resting-state functional brain networks have been used previously to identify the EZ [11, 27].

In general, most of the studies cited above made use of highly detailed non-linear models, such as neural mass models or theta models [49]. These models depend on several parameters that need to be adjusted beforehand, which complicates the optimization of the model and makes it difficult to obtain conclusions that are generalizable. As it is usually the case, an interplay exists between the generalizability and accuracy of the model, such that optimizing the predictive power of a model often means reducing the number of tunable parameters [50]. Thus, simpler models with few parameters might prove more reproducible, especially if the behavior of the model is understood mathematically. In this regard, one particular framework of relative mathematical simplicity that may capture the fundamental aspects of seizure propagation is that of epidemic spreading models [51]. These models simulate the propagation of an agent from some given location to other connected areas, a basic phenomenon appearing in a multitude of systems. Such models have been used, for instance, to study the spreading of pathological proteins on brain networks [52], or the relation between brain structure and function [53].

We propose that epidemic models can also provide a good representation of the initial steps of seizure propagation, during which the anomalous highly synchronized ictal activity propagates from the EZ to other regions. Moreover, as it is the case in epilepsy surgery, studies on epidemic models often try to find ways to stop or limit the propagation of the epidemics. Thus, epidemic models could also aid the planning of epilepsy surgery. For instance, the fundamental role of hubs in propagation is a well-known result of epidemic spreading [54], and targeting hub regions is often the most efficient way to obtain global immunization [55, 56]. Data of outbreak patterns can also be used retroactively to find the location of the origin of an epidemic [57, 58]. Within this framework, in a previous study [41] we modelled seizure propagation as an epidemic spreading process and, using the eigenvector centrality as a surrogate measure, found that the size of the resection area could be largely reduced with only a small decrease in the efficacy of the virtual surgery.

Here we defined an individualized seizure propagation model by making use of the Susceptible-Infectious (SI) model of epidemic spreading on top of a global brain network, for a group of 10 epilepsy patients who underwent epilepsy surgery, and for whom the surgical outcome at least one year after surgery was known. The network was based on the patient-specific MEG functional connectivity. The model parameters were tuned using information about the patient-specific seizure propagation patterns in invasive EEG recordings and the location of the resection area. First, we showed that, despite its simplicity, the model provides a good approximation to clinically observed seizure propagation patterns, and we fit the free parameters of the model to optimize the reproduction of the individual seizures. Secondly, we used the individualized model to test alternative resection strategies and, making use of an optimization algorithm, we found optimal – in terms of reduction of seizure propagation – personalized resections.

## 2 Methods

### 2.1 Patient group

We retrospectively analyzed 10 patients (5 females) with refractory epilepsy (Table 1). All patients underwent epilepsy surgery at the Amsterdam University Medical Center, location VUmc, between 2016 and 2019. All patients had received a magnetoencephalography (MEG) recording, had undergone a SEEG (stereo-electro-encephalography) study, and underwent pre- and post-surgical MR imaging. All patients gave written informed consent and the study was performed in accordance with the Declaration of Helsinki and approved by the VUmc Medical Ethics Committee.

**Table 1:**
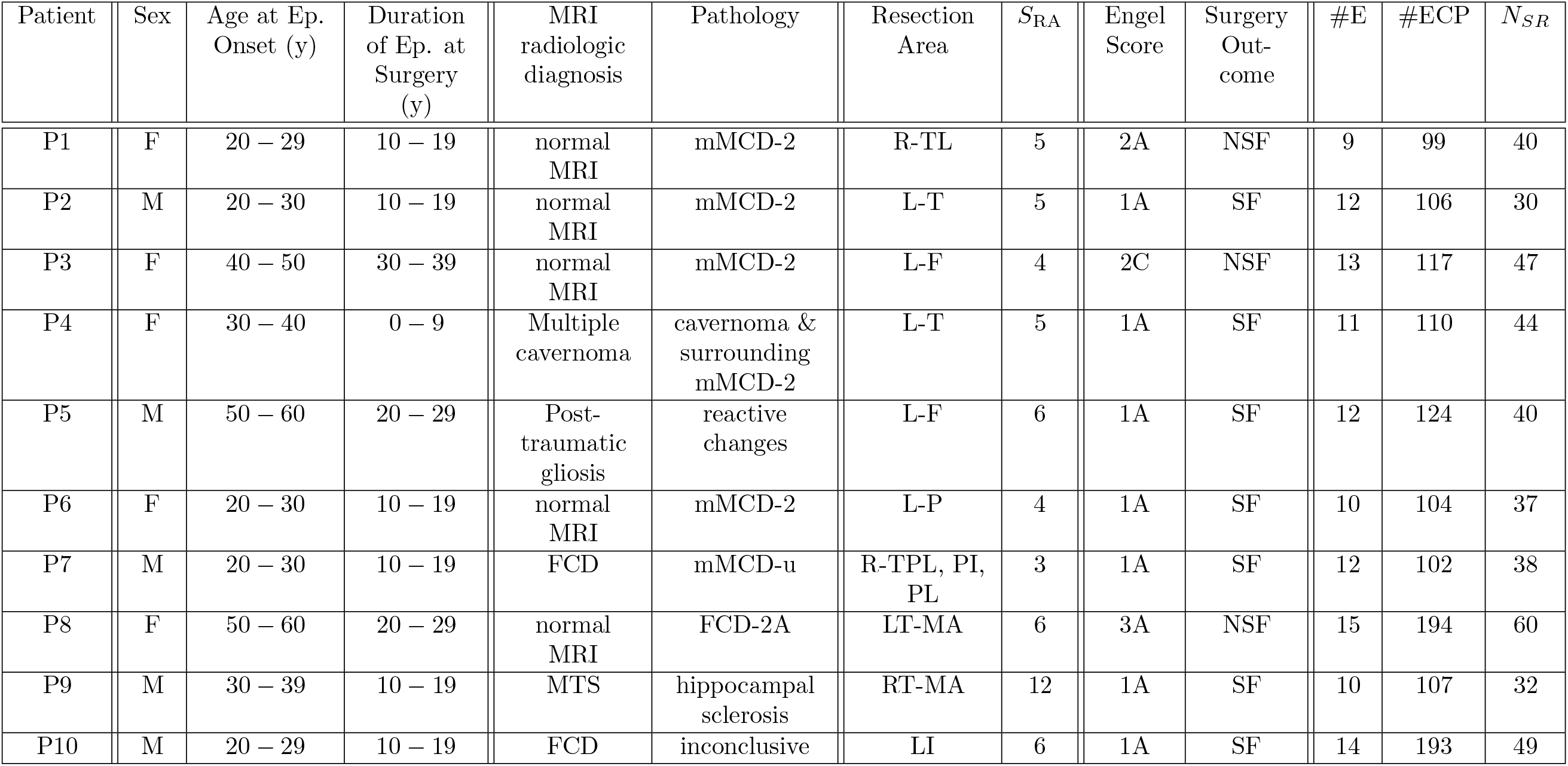
Patient data. Ep. = Epilepsy, y = years, *S*_RA_ = number of resected ROIs, #E = number of intracranial electrodes, #ECP = total number of electrode contact points, *N*_*SR*_ = number of BNA ROIs sampled by the SEEG electrodes. F = female, M = male, R = right, L = left, F = frontal, T = temporal, P = Parietal, TL = lateral temporal, TPL = posterior lateral temporal, PI = posterior insula, PL = posterior parietal, FCD = focal cortical dysplasia, mMCD = mild Malformation of Cortical Development, mMCD-2 = mMCD type 2, mMCD-u = mMCD type unknown, FCD-2A = focal cortical dysplasia type 2A, SF = seizure free, NSF = not seizure free.

The patient group was heterogeneous with temporal and extratemporal resection locations and different etiology. Surgical outcome was classified according to the Engel classification at least one year after the operation [59]. Patients with Engel class 1A were labelled as seizure free (SF), and patients with any other class were labelled as non seizure free (NSF). 3 patients were deemed NSF.

### 2.2 Individualized Brain Networks

The individualized computer model was based on the functional brain network of each patient (see figure 1), which was reconstructed in the Brainnetome Atlas (BNA) from MEG scans, as detailed below. Pre-operative Magnetic resonance imaging (MRI) scans were used for co-registration with the MEG data. MRI T1 scans were acquired on a 3T whole-body MR scanner (Discovery MR750, GE Healthcare, Milwaukee, Wisconsin, USA) using an eight-channel phased-array head coil. Anatomical 3D T1-weighted images were obtained with a fast spoiled gradient-recalled echo sequence. During reconstruction, images were interpolated to 1mm isotropic resolution.

**Figure 1:**
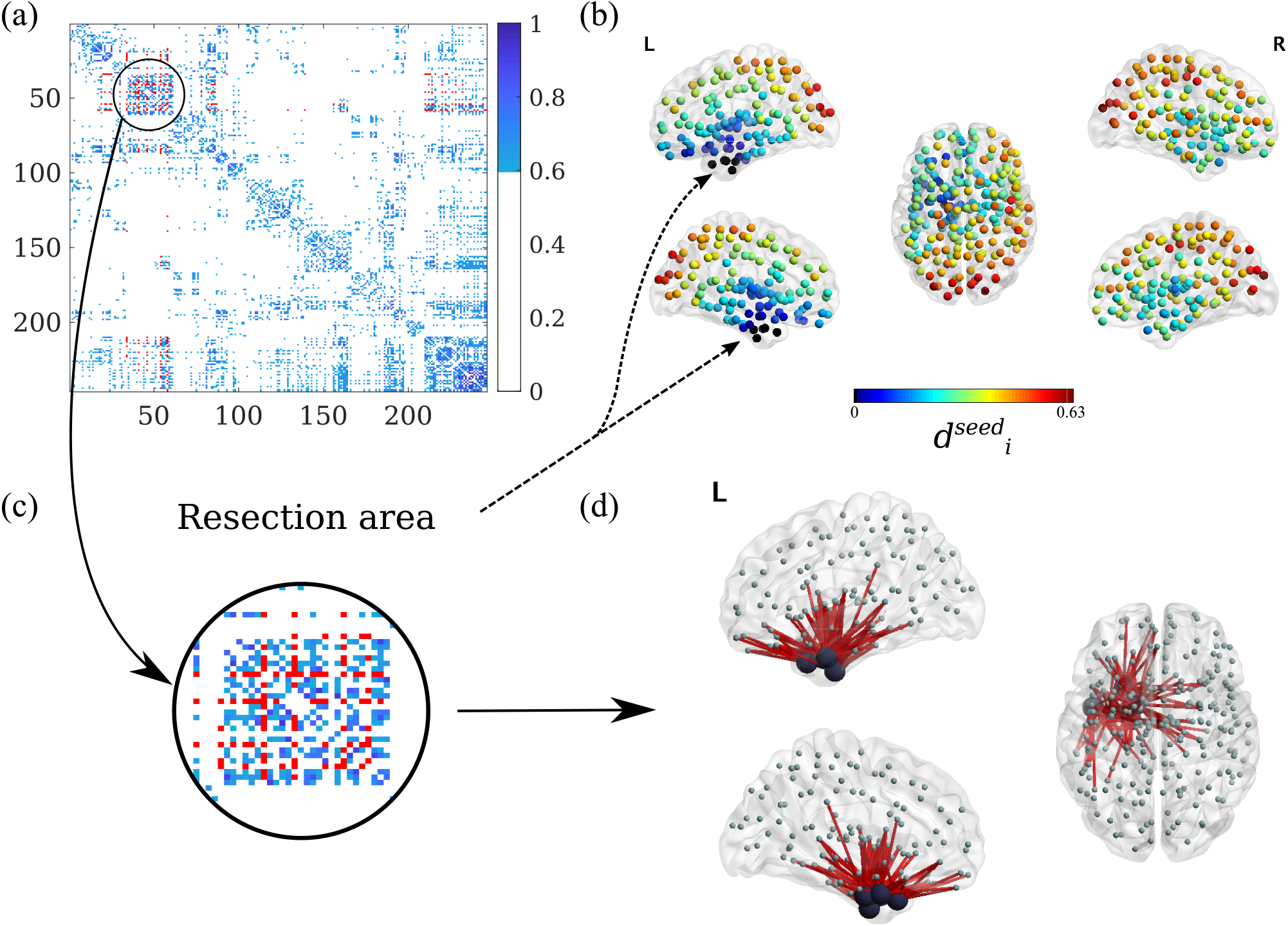
Individual Brain Networks. **a)** Weighted (and thresholded) resting-state broadband-MEG connectivity matrix for patient 4, for *θ* = 0.15. Each entry corresponds to a BNA region, and the regions have been re-ordered to group regions in the same hemisphere. In this representation, ROIs 1-105 correspond to the left cortical regions, ROIs 106-210 to the right cortical regions and ROIs 211-246 to the subcortical regions (in alternating hemisphere order). The strength of each connection is indicated by the colorcode. In red we show the connections from and to the resection area (RA). **b)** Distribution of average distances to the RA according to eq.(S2) for patient 4, in dimensionless units. The points mark the centroids of the BNA ROIs, and the color scale indicates the effective distance of the ROI to the resection area (RA), which appears as black dots. **c)** Zoom-in of the adjacency matrix: RA and surrounding nodes. **d)** Illustrative representation of the RA (big black circles) and all the links connecting it with the rest of the network. Figures with brain representations have been obtained with the BrainNet Viewer [68].

#### 2.2.1 MEG acquisition

MEG recordings were obtained during routine clinical practice using a whole-head MEG system (Elekta Neuromag Oy, Helsinki, Finalnd) with 306 channels consisting on 102 magnetometers and 204 gradiometers. The patients were in supine position inside a magnetically shielded room (Vacummshmelze GmbH, Hanau, Germany).

Typically, three data-sets of 10 to 15 minutes each containing eyes-closed resting-state recordings were acquired and used in the presurgical evaluation for the identification and localization of interictal epileptiform activity. Paradigms for the localization of eloquent cortex, such as voluntary movements and somatosensory stimulation [60], as well as a hyperventilation paradigm to provoke interictal epileptiform discharges, were also recorded but not analysed in this study. The data were sampled at 1250 Hz, and filtered with an anti-aliasing filter at 410 Hz and a high-pass filter of 0.1 Hz. The head’s position relative to the MEG sensors was determined using the signals from four or five head-localization coils that were recorded continuously. The positions of the head-localization coils and the outline of the scalp (roughly 500 points) were measured with a 3D digitizer (Fastrak, Polhemus, Colchester, VT, USA), and were later used for co-registration with the anatomical MRI.

The temporal extension of Signal Space Separation (tSSS) [61, 62] was used to remove artifacts using Maxfilter software (Elekta Neuromag, Oy; version 2.1). The points on the scalp surface were used for co-registration with the anatomical MRI of the patient through surface-matching software. A single sphere was fitted to the outline of the scalp and used as a volume conductor model for the beamforming approach. For a detailed description and parameter settings see [60].

#### 2.2.2 MEG processing: Atlas-based Beamforming

Neuronal activity was reconstructed using an atlas-based beamforming approach, modified from [63], in which the time-series of neuronal activation of the centroids of the ROIs were reconstructed [64]. We considered the 246 ROIs in the BNA atlas [65], including 36 subcortical ROIs, whose centroids were inversely transformed to the co-registered MRI of the patient. Then, a scalar beamformer (Elekta Neuromag Oy; beamformer; version 2.2.10) was applied to reconstruct each centroid’s time-series. The beamformer weights were calculated for each centroid separately to form a spatial filter so as to maximally let pass signals that originate from the centroid of interest and to attenuate all other signals. The weights were based on the lead fields (using the spherical head model and an equivalent current dipole as source model), the data covariance and noise covariance. The broadband (0.5 – 48.0 Hz) data covariance was based on the entire recording (on average 799.23 seconds of data (range: 309.50 - 908.67) were used). A unity matrix was used as noise covariance when estimating the optimum source orientation for the beamformer weights [66]. The broadband data were projected through the normalised beamformer weights [67] in order to obtain time-series (virtual electrodes, VE) for each centroid [64].

#### 2.2.3 Brain Functional Networks

The time-series for each VE were visually inspected for epileptiform activity and artifacts. On average, 53.1 (range: 19 – 60) interictal and artefact-free epochs of 16384 samples were selected for each patient. The epochs were further analyzed in Brainwave (version 0.9.151.5 [69]) and were down-sampled to 312 Hz, both in the broadband (0.5 - 48 Hz) and in the alpha-band (8 - 13 Hz).

Functional networks were generated using the 246 VEs as nodes (see figure 1a). Functional connectivity (i.e. the elements *w*_*ij*_ of the weight matrices) was estimated by the AEC (Amplitude Envelop Correlation) [70–73]. The uncorrected AEC (i.e. without correcting for volume conduction) connectivity metric was selected as it maintains information on the structural connectivity pattern, whilst including information on long-range functional connections. AEC values range from 0 (no functional connectivity) to 1 [74]. Functional networks were thresholded at different levels *θ* indicating the percentage of remaining links, and the resulting average connectivity *κ* = *θN* of the network was determined. We considered a non-uniform grid in *θ*, with values *θ* = 0.01, 0.02, 0.04 …, 0.10, 0.15, 0.50, to account for the fact that the model is more sensitive to connectivity changes for small *θ*. Notice that the networks were thresholded but not binerized, so that *w*_*ij*_ remains a real variable (*w*_*ij*_ ∈ [0, 1]). The resulting weight matrix is represented in figure 1a for a characteristic case.

#### 2.2.4 Resection Area

The resection area (RA) was determined for each patient from the three-month post-operative MRI. This was co-registered to the pre-operative MRI (used for the MEG co-registration) using FSL FLIRT (version 4.1.6) 12 parameter affine transformation. The resection area was then visually identified and assigned to the corresponding BNA ROIs, name those that were overlapping for at least 50% with the resection area. In figure 1 we illustrate the resection area and its connectivity structure with the rest of the network for one patient.

### 2.3 Individualized Propagation Pattern

All patients underwent stereo-electroencephalography (SEEG) electrode implantation. The number and location of the intracerebral electrodes (Ad-Tech, Medical Instrument Corporation, USA, 10-15 contacts, 1.12 mm electrode diameter, 5 mm intercontact spacing; and DIXIE, 10-19 contacts, 0.8 mm electrode diameter, 2 mm contact length, 1.5 insulator length, 16 – 80.5 insulator spacer length) were planned individually for each patient by the clinical team, based on the location of the hypothesized seizure onset zone (SOZ) and seizure propagation pattern. Implantation was performed with a stereotactic procedure. The number of electrodes per patient varied between 9 and 15 (average = 11.8) and the total number of contacts between 99 and 194 (average = 125.6). Details of the number of electrodes and contact points for each patient are indicated in table 1.

The locations of the SEEG contact points were obtained from the post-implantation CT scan (containing the SEEG electrodes) that was co-registered to the preoperative MRI scan using FSL FLIRT (version 4.1.6) 12 parameter afine transformation (see figure 2*a*). Each electrode contact point (CP) was assigned the location of the nearest ROI mass center. Because BNA ROIs are in general larger than the separation between contact points, different CPs can have the same assigned ROI. We refer to the set of ROIs sampled by the SEEG CPs as *SEEG*_ROI_, with the size of the set being *N*_*SR*_ (*N*_*SR*_ values for all patients are reported in table 1).

**Figure 2:**
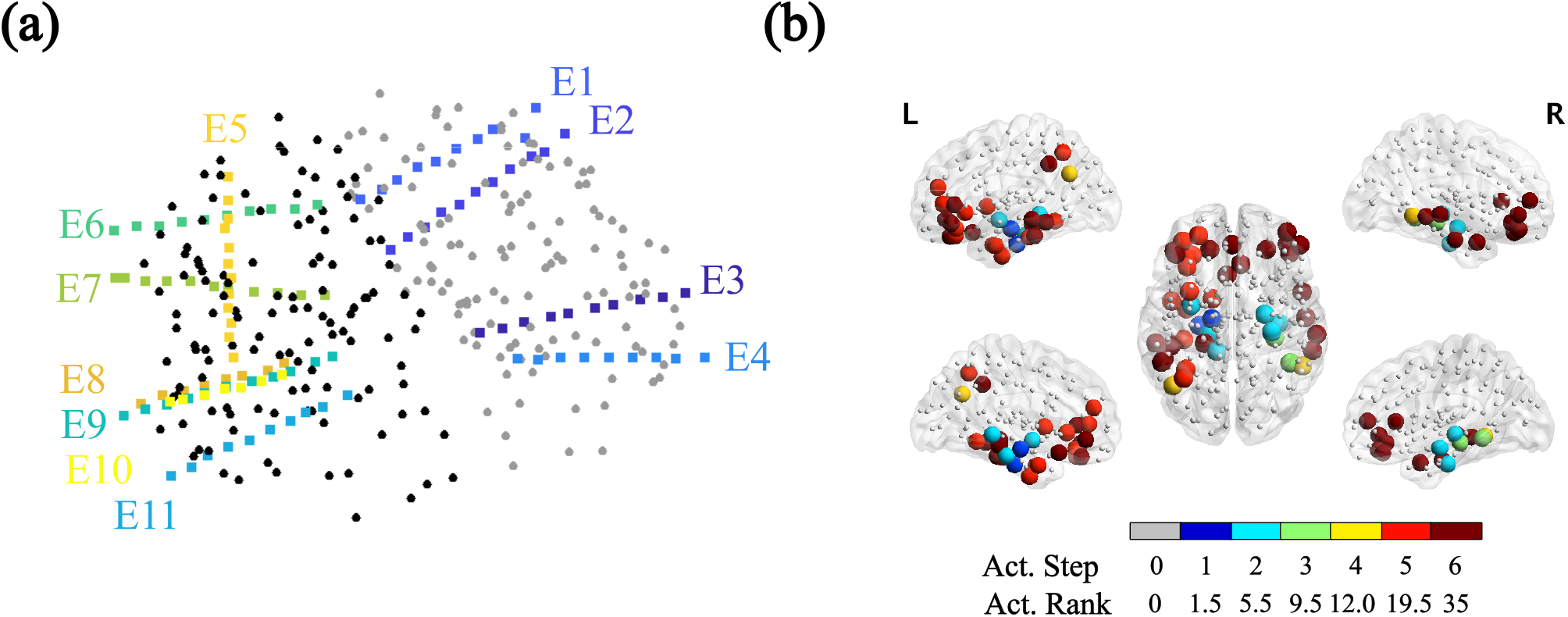
Individualized Seizure Propagation Pattern. **a)** SEEG electrodes for patient 4. Black and gray dots indicate the BNA ROIs’ mass centers, respectively for the left and right hemispheres (different colors are used for visualization purposes). This patient had 11 intracranial electrodes implanted, each electrode is shown in a different color. **b)** Corresponding seizure pattern constructed from the clinical SEEG recordings. First, different activation steps were identified in the seizure recordings, together with the corresponding contact points (CPs). In the case depicted here, typical seizures consisted of 6 propagation steps, with step 1 depicting the seizure onset zone (as indicated by the SEEG recordings), and step 6 signalling the generalization of the seizure to all sampled CPs (note however that in general not all CPs need take part in the seizure propagation pattern). This propagation pattern was then translated into the BNA space, and the sampled ROIs are indicated here as large colored spheres. Thus, only the BNA ROIs sampled by the SEEG electrodes are included in the pattern, which in this case corresponded to a total of *N*_*RS*_ = 49 sampled ROIs (*N*_*RS*_ values for all patients are reported in table 1). The color code in the figure indicates the propagation step in which the corresponding CP is involved in ictal activity (i.e. Act. Step). Small grey dots mark the ROIs not included in the SEEG pattern. Finally, in order to enable comparison with the SEEG propagation pattern with the one modelled via the SI dynamics via the Mann-Whitney U test, we calculated the activation rank (Act. Rank) of each ROI, such that the ROIs were ordered and ranked according to the activation step, and groups with the same rank were assigned a rank equal to the midpoint of unadjusted values.

Based on the clinical recordings, a seizure propagation pattern was built indicating the order of activation of the electrode CPs for a typical seizure, as shown in figure 2*b*. In order to do so, the start of ictal activity of typical seizures was visually assessed for each SEEG CP by a clinician expert. Then, the CPs were grouped into activation steps according to when ictal activity was first observed. The seizure pattern was built from one typical seizure for each patient. This activation pattern was then translated into the BNA space (see figure 2*b*), so that the each ROI *i* in the sampled set SEEG_ROI_ was assigned an activation step. Finally, we calculated the *activation rank* of each ROI in SEEG_ROI_, such that the SEEG_ROI_ ROIs were ordered and ranked according to their activation step, and groups with the same rank (i.e. in the same activation step) were assigned a rank equal to the midpoint of unadjusted values (see figure 2b). This yields the SEEG pattern 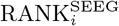 indicating the activation rank of each ROI *i* in SEEG_ROI_.

### 2.4 Seizure Propagation Model

#### 2.4.1 SI Dynamics

Seizure propagation was modelled via an epidemic spreading model: the susceptible-infected (SI) model [51]. This model depicts the propagation of an epidemic process on a network from a set of seed regions to the rest of the nodes. The model only accounts for the propagation process, as it does not include any mechanisms for the deactivation of the affected regions. It thus represents the initial steps of the propagation of a seizure, before inhibition takes place and the affected regions start to deactivate. Thus, we do not try to mimic the complicated processes involved in seizure generation and propagation with this model, which is used here as an abstraction that includes only the most relevant features of the initial steps of seizure propagation. Thanks to its simplicity, the model can be described by using only one parameter, the infection probability *β*, as described below. More complicated epidemic models, such as the SIR or SIS model [51], which include a deactivation mechanism, introduce more parameters that either have to be assumed or fitted with detailed data.

Simulation of the epidemics on the network takes place as follows. Each node is characterized by its state: either S (susceptible) or I (infected). Initially, all nodes are in the S state, except for a set of nodes in the I state, which act as the seed of the epidemic (or seizure). At each time step, each infected node can propagate the infection independently to any of its neighbours with probability *βw*_*ij*_, where *β* characterizes the rate of the epidemic spreading and *w*_*ij*_ is the connection strength between nodes *i* and *j* as given by the MEG-FC adjacency matrix. The fraction of infected nodes at each time is given by *I*(*t*). If all nodes are connected, eventually the epidemic spreads over the whole network, *I*(*t* → ∞) = 1, as shown in figure 3a. However, the speed and pattern of the propagation depend on *β* and *w*_*ij*_ (see figure 3).

**Figure 3:**
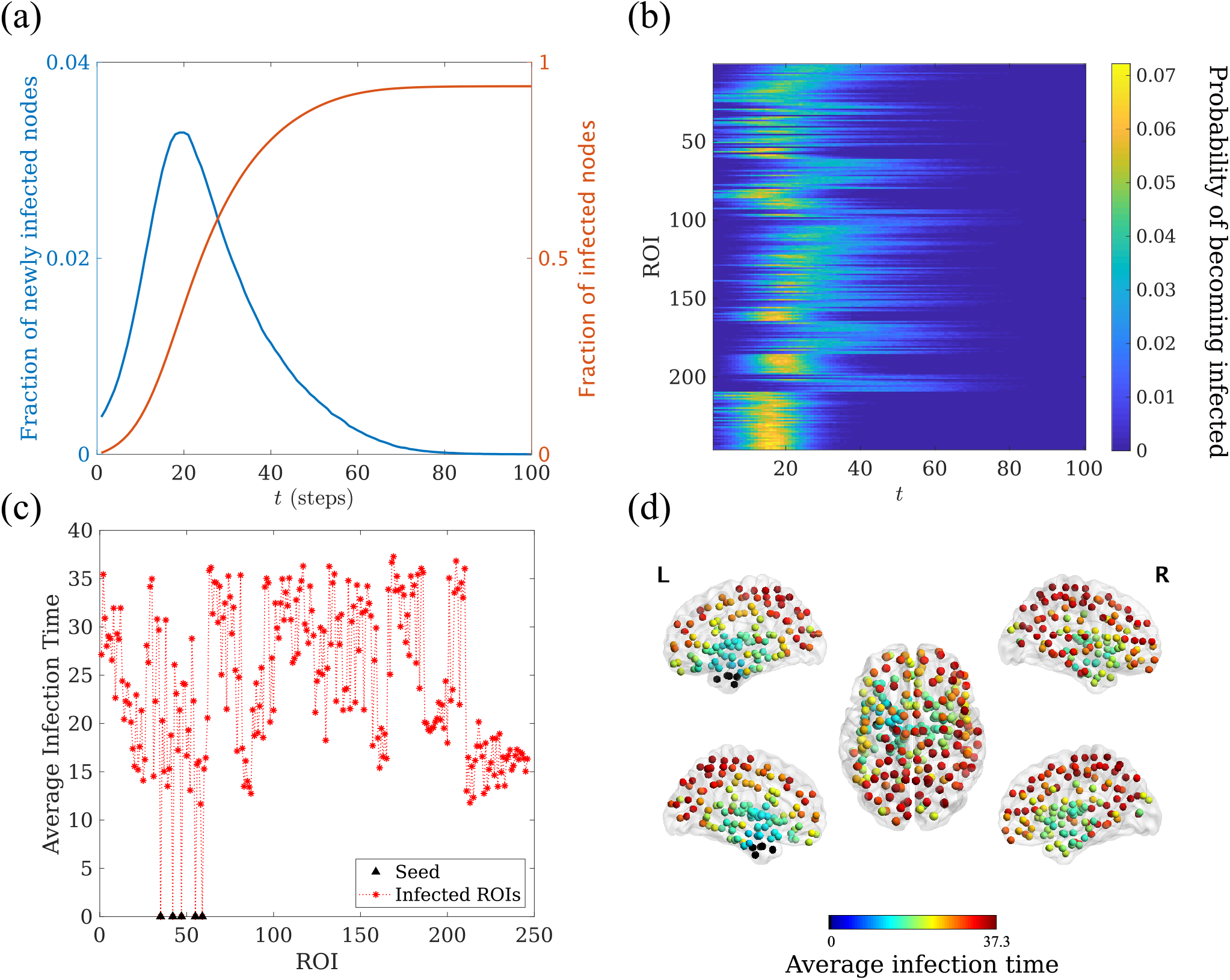
Seizure propagation model. Exemplary SI propagation process for patient 4. **(a)** *I*(*t*) (red line) and the fraction of newly infected nodes at time *t* (*I*(*t*) *−I*(*t −*1), blue line), as functions of time. **(b)** Model propagation pattern showing the probability *p*_*i*_(*t*), as indicated by the color scale, that a given ROI *i* (y-axis) becomes infected at time *t* (x-axis). **(c)** Average infection time for each ROI, *t*_*i*_. The seed (corresponding to the resection area, shown in black triangles) is always infected at time 0. **(d)** Spatial representation of the ROIs mean infection time *t*_*i*_. Each ROI is color-coded according to its average infection time. The resection area is shown as black circles. The time unit is the number of simulation steps in all panels.

In order to fit and validate the model, we first considered the situation of slow propagation in which only one new node is infected at each time step (formally corresponding to *β* → 0), and compared the propagation pattern of the modelled epidemic process with the clinical SEEG seizure pattern for different connectivity thresholds *θ*. The threshold was then fit to maximize the correlation between the modelled and clinical propagation patterns. Then, to study the effect of different virtual resections, we quantified the short-term propagation of the seizure as the fraction of infected nodes at time *t*_0_. Here we set *t*_0_ = 50 and, in order to account for different network densities, we set *βθ* = const = 4 · 10^*−*4^ (so that *β* = 0.01 for a network with 4% of the links, for instance). For a standard resection size *S*_RA_ of 4 nodes this would correspond, in the uniform limit, to an infection of about 2*/*3 of the nodes [50].

SI dynamics was simulated in custom-made Matlab algorithms using Monte-Carlo methods, with *N*_*R*_ = 10^4^ iterations of the algorithm for each configuration to assure convergence.

#### 2.4.2 Optimization of SI parameters: Individualized Propagation Model

The SI dynamics were simulated as described above, leading to a probability map indicating the probability *p*_*i*_(*t*) that each ROI *i* became infected at step *t*, for each connectivity threshold *θ*, as shown in figure 3*b*. The mean activation time for each ROI was then calculated as 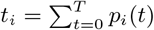, where *T* is the maximum integration time. *t*_*i*_ describes the activation sequence of the ROIs during a modelled seizure (see figure 3*c, d*). Given that not all BNA ROIs were sampled by the SEEG electrodes, *t*_*i*_ was then sub-sampled to the SEEG_ROI_ set, and the included ROIs were ranked according to *t*_*i*_. This ranking 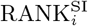 constitutes the modelled or SI seizure propagation pattern, which is defined upon the same set of ROIs SEEG_ROI_ as the clinical one 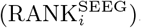, as shown in figure 4.

**Figure 4:**
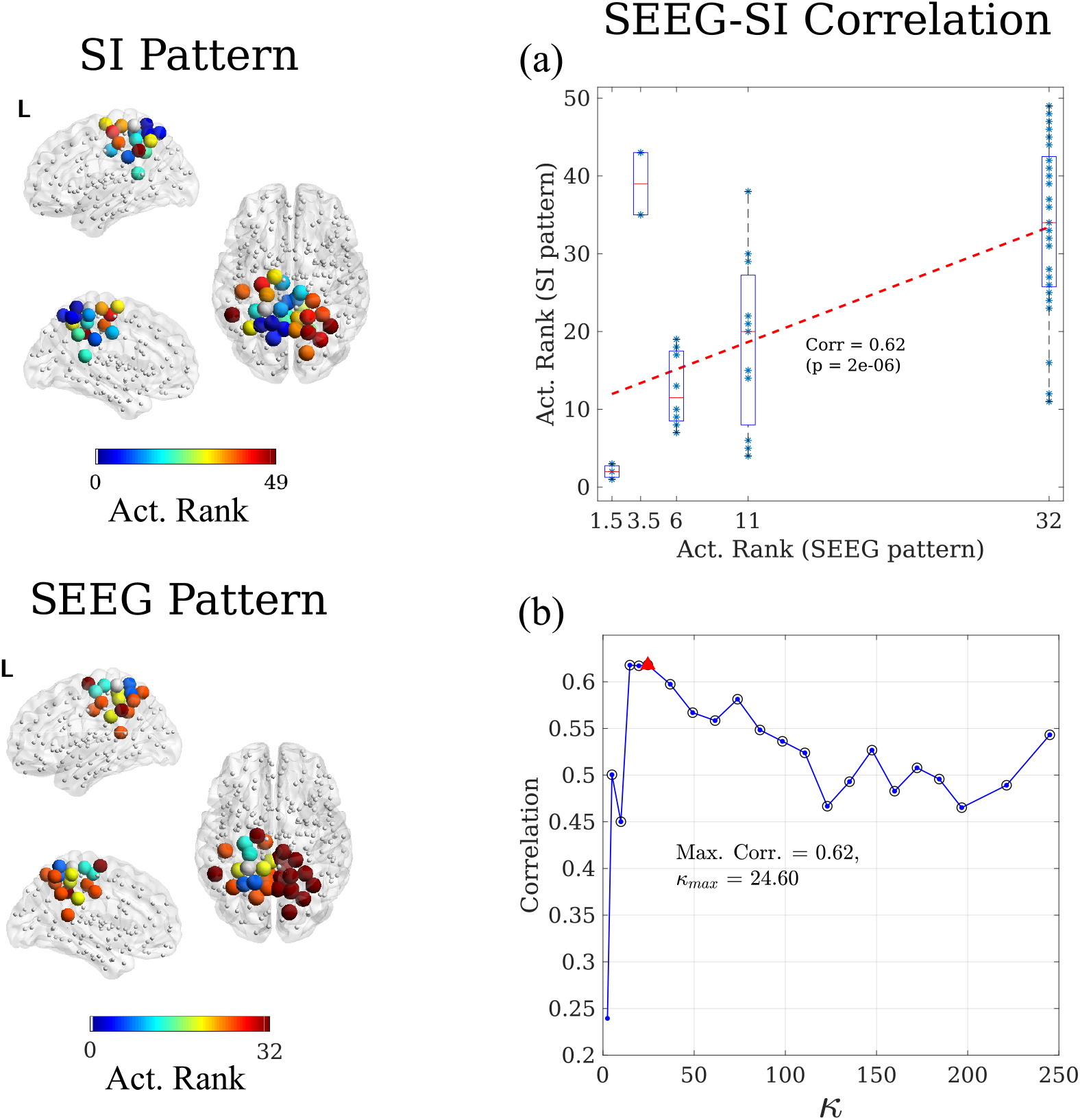
Correlation method. Here we illustrate, for patient 3, the correlation method to validate and fit the seizure propagation model. For each network connectivity threshold *θ*, the SI dynamics was simulated over the whole MEG network, and the propagation pattern was constructed for the ROIs sampled by the SEEG electrodes and compared with the clinical SEEG pattern. Each pattern describes the activation order – or rank – of each sampled ROI. Given that the clinical SEEG pattern is built in term of activation steps, different ROIs can have the same ranking, as described in the main text. The modelled SI pattern was correlated with the clinical SEEG pattern, as depicted in panel **(a)**. This process was iterated for different connectivity thresholds *θ* leading the correlation curve shown in panel **(b)**, where the mean degree *κ* = *θN* is shown in the x-axis, and significant correlations are indicated with a black circle. Finally, the connectivity leading to the maximum correlation was chosen. In the depicted case, this corresponded to a mean degree of *κ* = 24.60 (*θ* = 0.1) leading a correlation of 0.62 (red triangle). This also corresponds to the value of *κ* used for panel a.

Once the SI pattern had been constructed, the ranked correlation was computed to compare the SI and SEEG patterns (see figure 4a) via a Mann-Whitney U test. The correlation is thus defined as

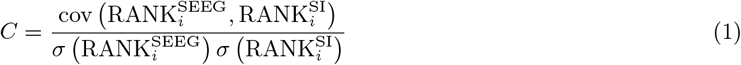

where cov(*x, y*) and *σ*(*x*) respectively stand for the covariance and standard deviation. The connectivity threshold that maximized this correlation was independently found for each patient (see figure 4b) and used for the corresponding individualized virtual resection model.

Two possible seeds were considered for the SI dynamics: either the resection area (RA seed), or the hypothesized Seizure Onset Zone according to the SEEG clinical recordings (SOZ seed).

### 2.5 Simulation of Resections

Resections ℛ of sets of nodes were conducted in the model by the use of *virtual* resections (VRs). For this, all the connections of the corresponding nodes were set to 0, so the size of the network was left unchanged, but the resected nodes became isolated. Each resection ℛ was then characterized by measuring the fraction of infected nodes at a fixed time *t*_0_ after the resection had taken place, *I*_ℛ_(*t*_0_) (see figure 5).

**Figure 5:**
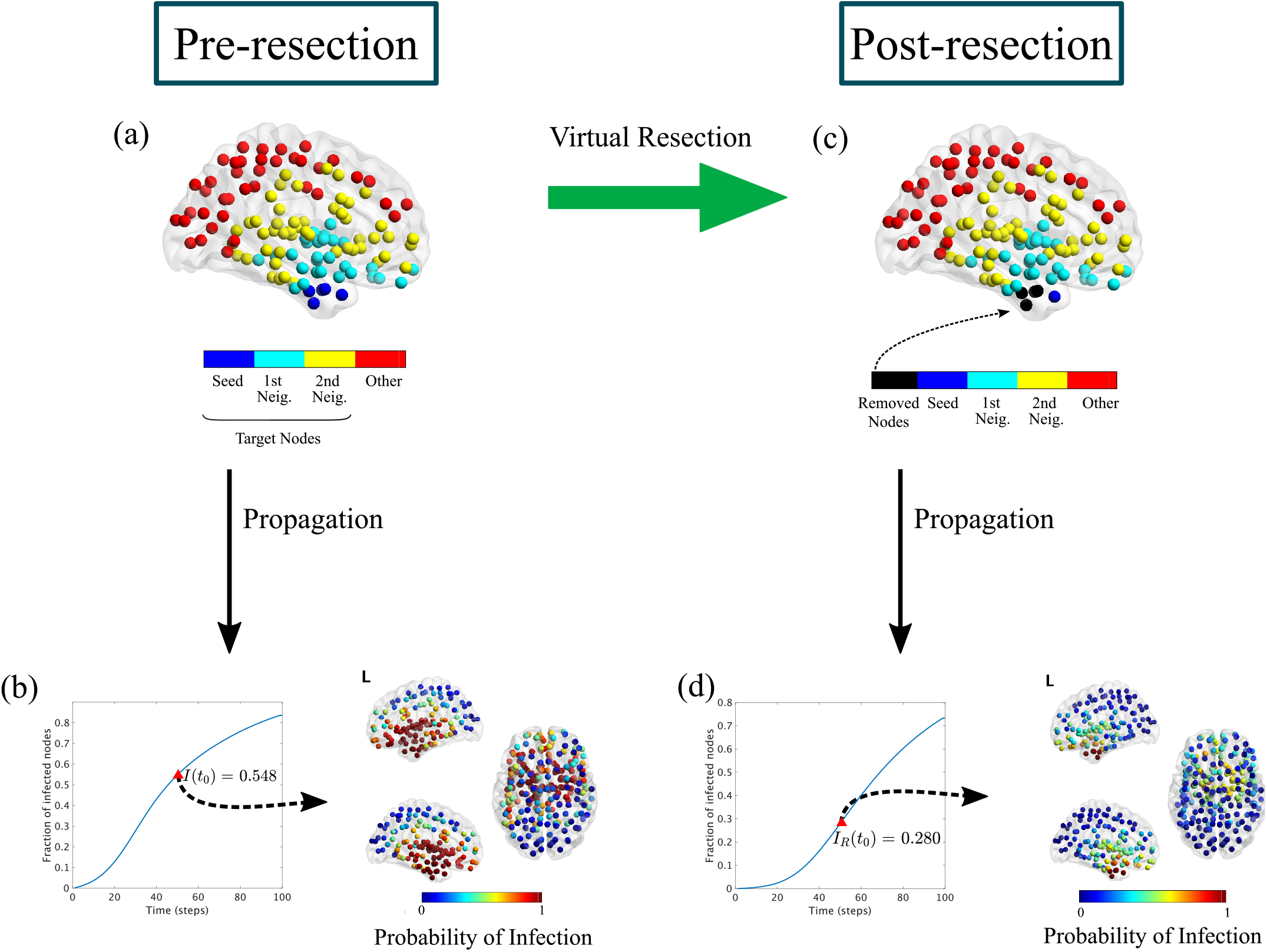
Virtual Resection Implementation. The target nodes for the virtual resection VR are all nodes at two steps or less from the seed, i.e. the seed and its first and second neighbours (panel **a**). The initial seizure propagation is the fraction of infected nodes at *t*_0_ = 50, *I*(*t*_0_) = 0.548 (panel **b**). A virtual resection of 5 nodes is implemented in the network by setting to 0 all the links with the corresponding nodes, marked in black in panel **c**. Seizure propagation is now reduced by approximately a factor 2 (*I*_*R*_(*t*_0_) = 0.280), and the probability that the seizure reaches regions outside the seed decreases considerably (panel **d**). This example corresponds to patient 4.

The goal of epilepsy surgery is to completely stop seizure propagation (*I*_ℛ_(*t*_0_) → 0 ∀*t*_0_) which, in the present model, can only be attained by (and it is always attained by [75, 76]) complete disconnection of the assumed seed region. Thus, in this study the effect of each resection was measured in terms of the decrease in the speed of seizure propagation, and the goal was to find the smallest resection able to reduce the initial propagation at *t*_0_ by 90%. That is, to find the smallest resection such that *i*_ℛ_ = *I*_ℛ_(*t*_0_)*/I*(*t*_0_) ≤ 0.1, where *I*(*t*_0_) is measured on the original pre-resection network [41].

We defined a four-step method to find the optimal resection ℛ*\** for each resection size *S*, ℛ*\**(*S*). That is, the resection leading to a minimum propagation *I*_ℛ*(*S*)_(*t*_0_). The optimization method made use of the Simulated Annealing algorithm [77] to speed up the exploration of the space of possible resections, and it considered a surrogate structural metric – the mean *effective distance* to the seed [50, 57, 58] – as a proxy for the SI dynamics to simplify the initial exploration (the method and algorithms used are described in detail in the Supplementary Information and figure S1). Then, the smallest resection leading to a 90% reduction in seizure propagation (as measured by *I*_ℛ_(*t*_0_)), ℛ_90_, was identified. We also identified, for each patient, the smallest resection leading to 100% reduction in propagation (*I*_ℛ_(*t*_0_) = 0), ℛ_100_. Finally, to characterize the effect of small resections in seizure propagation, we also defined the one-node resection, ℛ1, as the resection of size 1 with a maximum effect.

In principle all nodes in the network could be considered as possible targets to be resected. However, the effect of each node on seizure propagation decreases as it gets further from the seed (in terms of hops on the network). Therefore, here we considered only nodes that were at most two hops (without taking into account the edge weights) away from the seed (that is, the seed and its first and second neighbours), as depicted in figure 5a.

### 2.6 Statistics

The Mann-Whitney U test was used to determine the correlation between the modelled and clinical seizure propagation patterns. To compare the optimal correlation obtained with different network definitions we used a paired Student’s t-test. Similarly, different seed definitions were compared using a paired Student’s t-test. Finally, for comparisons between SF and NSF patients, we used an unpaired Student’s t-test. All significance thresholds were set at *p <* 0.05.

Finally, analyzed the effect of the size and mean degree of the resection area on the size of the 90% and 100% resections, and on the effect of the one-node resection, with a linear least squares fit.

### 2.7 Data availability

The data used for this manuscript are not publicly available because the patients did not consent for the sharing of their clinically obtained data. Requests to access to the datasets should be directed to the corresponding author. All user-developed codes are available from the corresponding author upon reasonable request.

## 3 Results

### 3.1 Preliminary results

A total of 10 patients (5 females) were included in the study, 7 of whom were deemed SF one year after surgery (see table 1 for the patient details).

We obtained the individual weighted MEG functional connectivity networks, with entries *w*_*ij*_ characterizing the coupling strength between ROIs *i* and *j*, for each patient in the alpha-band and the broad-band using the BNA atlas (246 nodes). The tables and figures shown in the main text report the results for the broad-band networks, details for the alpha-band can be found in the Supplementary Information. An exemplary FC matrix is shown in figure 1.

We constructed a clinical propagation pattern from the seizures observed with SEEG, for each patient. The pattern was initially defined in term of the electrodes CP, and then translated into the ROIs of the BNA atlas (see figure 2).

### 3.2 Reproduction of Seizure Propagation Patterns

As described in the methods section (see figure 4), we estimated the correlation *C* between the SI seizure pattern and the SEEG pattern for a range of connectivity values *κ*, as shown in figure 6a for the broadband networks and in figure S3 for the alpha-band networks. Then, to fit the spreading model to the SEEG propagation data, we selected the connectivity value *κ*_max_ that maximized *C*(*κ*), for each patient. The maximum correlation *C*_max_ obtained for each patient and the corresponding *κ*_max_ are shown in figure 7*a, b* for the broadband networks and in figure S4 for the alpha-band networks, and the corresponding values are reported in tables S1 and S2, respectively. Most cases presented a bimodal dependence of the correlation on the network density, so that there was a maximum for low density and another maximum for large density. Here we restrict our analyses to the first maximum, which yields only the fundamental connections that are needed to reproduce seizure propagation.

**Figure 6:**
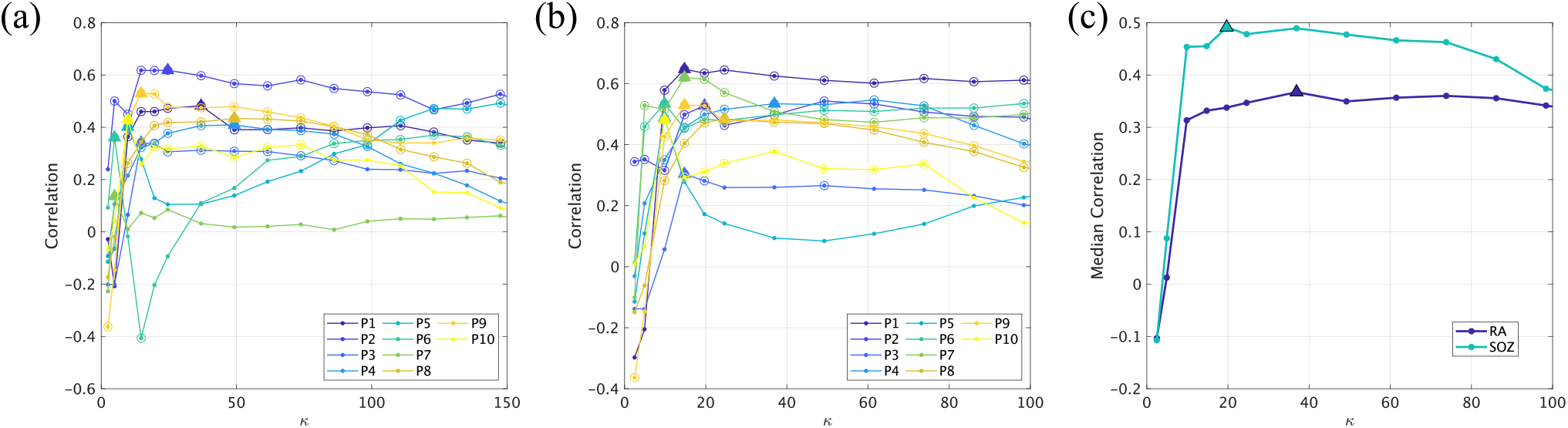
Reproduction of seizure propagation. Correlation between the modelled and clinically observed seizure propagation patterns as a function of network density, for each patient, using the RA (panel **a**) and SOZ seeds (panel **b**). Panel (**c)**) indicates the median curves for each case, as indicated by the legend. Circles in panels a and b denote significant correlations (*p <* 0.05), and the optimal correlation for each case is marked with a triangle.

**Figure 7:**
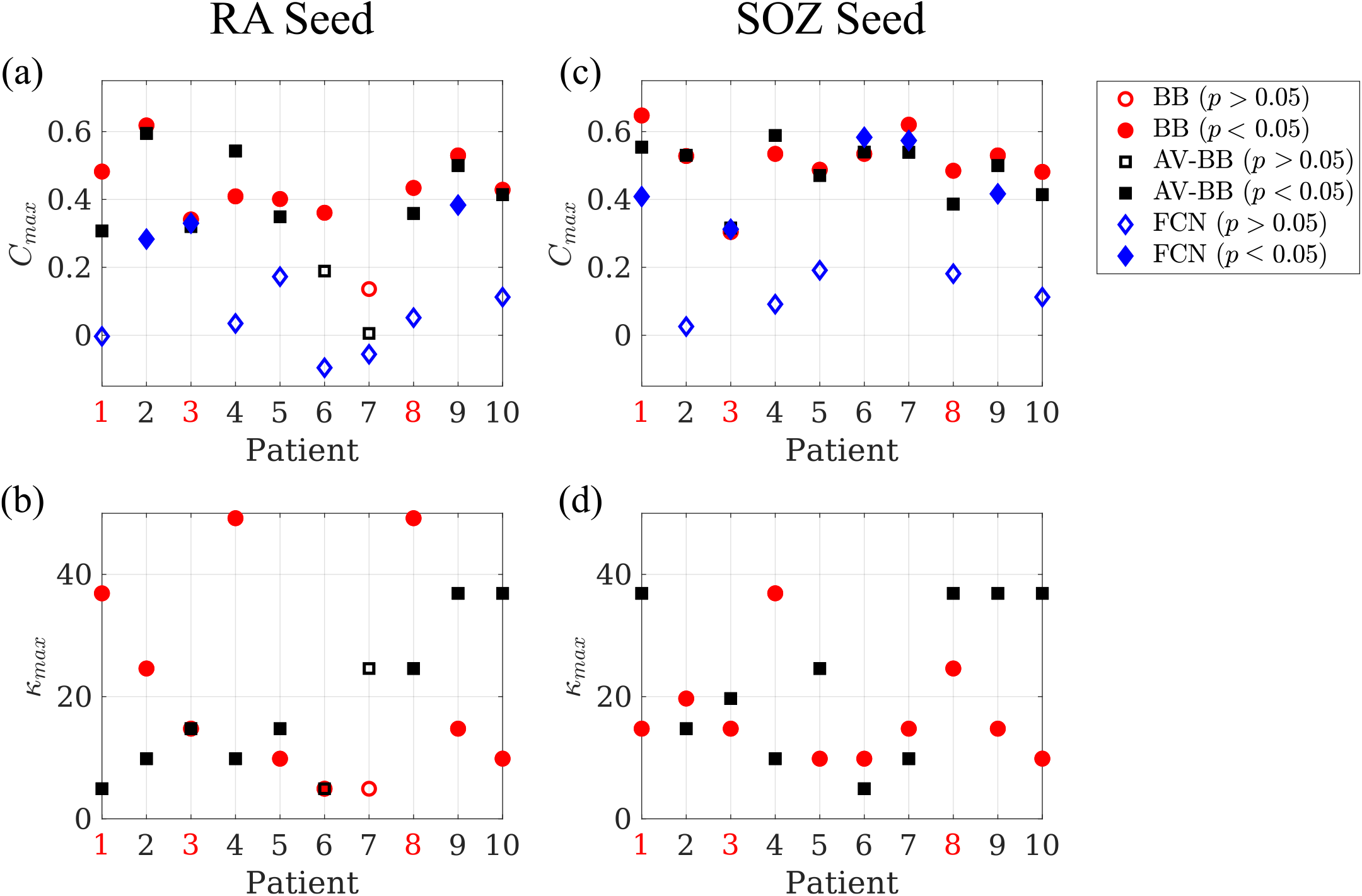
Reproduction of seizure propagation patterns. Panels **(a)** and **(c)** show the average maximum correlation *C*_*max*_ achieved by the individual BB networks (BB, red circles), and the average BB one (AV-BB, black squares), and the correlation found for the fully connected network (FCN, blue diamonds), respectively for the RA and SOZ seeds. Panels **(b)** and **(d)** show the corresponding *κ*_max_ for the individual (BB, red circles) and average (AV-BB, black squares) networks. Significant correlations (*p <* 0.05) are indicated by a filled marker, and non-significant ones (*p >* 0.05) by an empty marker. NSF patients are indicated by red labels.

We found that the model significantly reproduced the seizure propagation patterns for 9*/*10 patients. The average correlation was *C*^*α*^ = 0.38 for the alpha-band (*α*) networks and *C*^BB^ = 0.41 for the broad-band (BB) networks. The difference between the two settings (0.03, BB *> α*) was not significant (*t*(9) = 1.81, *p* = 0.06). There were no significant differences in the optimal correlation between SF and NSF patients (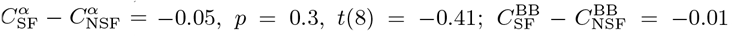, *p* = 0.5, *t*(8) = −0.07, unpaired Student t-tests). The optimal network density did not differ significantly between frequency bands (*κ*^BB^ − *κ*^*α*^ = 0.99, *p* = 0.4, *t*(9) = 0.15) or between the sub-groups(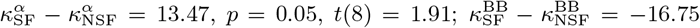, *p* = 0.09, *t*(8) = −1.50).

#### 3.2.1 Alternative Definition of the Seed

Different definitions of the seed can be considered. So far, we used the resection area, but in prospective studies the actual resection area will not be known. We therefore also considered the SOZ, as defined by the SEEG study, as the seed for the SI spreading (seed SOZ), and repeated the fit method as before (see figure 6b). We found that now the correlation between the model and the seizure pattern was significant for all patients, both for the alpha- and broad-band networks. The average correlations were respectively *C*^*α*^ = 0.47 and *C*^BB^ = 0.51. The difference (*C*^*α*^ − *C*^BB^ = −0.04) was significant (*p* = 0.04, *t*(9) = −1.99).

We found that the optimal correlation was higher for SF than for NSF patients 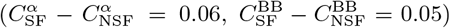, although the difference was not significant (for *α*-networks: *p* = 0.2, *t*(8) = 0.79; for BB networks: *p* = 0.2, *t*(8) = 0.79). The optimal network density did not differ significantly between frequency bands (*κ*^BB^ − *κ*^*α*^ = −6.64, *p* = 0.08, *t*(9) = −1.52) or between the sub-groups (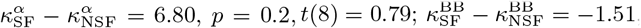 *p* = 0.4, *t*(8) = −0.2).

Overall, this seed definition reproduced the clinical seizure pattern better than the RA seed (*C*^*α*^(SOZ) − *C*^*α*^(RA) = 0.09, *p* = 0.07, *t*(9) = 1.63; *C*^BB^(SOZ) − *C*^BB^(RA) = 0.10, *p* = 0.04, *t*(9) = 1.99), although the difference was only significant for broad-band networks.

#### 3.2.2 Effect of Individualized Brain Networks

Is patient specific connectivity required to reproduce the clinically observed seizure propagation patterns? In order to answer this question, we repeated the analysis using the average functional connectivity matrix (referred to as AV-*α* and AV-BB respectively for the alpha-band and broadband networks) as the network backbone for the SI spreading dynamics for all patients.

For the broad-band network, a significant correlation was found for 8 (10) patients using the RA (SOZ) seed (see figures 7 and S4). The average optimal correlation (*C*^AV-BB^(RA) = 0.36, *C*^AV-*α*^(SOZ) = 0.48) was smaller than for the individual patient networks (*C*^BB^(RA) − *C*^AV-BB^(RA) = 0.05, *p* = 0.08, *t*(9) = 1.95; *C*^BB^(SOZ) − *C*^AV-BB^(SOZ) = 0.03, *p* = 0.09, *t*(9) = 1.92), although the difference was not significant. For the alpha-band network, a significant correlation was found for 7 (10) patients using the RA (SOZ) seed. The average (optimal) correlation (*C*^AV-*α*^(RA) = 0.34, *C*^AV-*α*^(SOZ) = 0.46) was smaller than for the individual patient networks (*C*^*α*^(RA) − *C*^AV-*α*^(RA) = 0.05, *p* = 0.2, *t*(9) = 1.30; *C*^*α*^(SOZ) − *C*^AV-*α*^(SOZ) = 0.01, *p* = 0.8, *t*(9) = 0.28) but the difference was not significant.

#### 3.2.3 Average model

Above, the optimal network connectivity was fitted independently for each patient using the SEEG data. In order to test if a mean model could be used for patients without SEEG recordings, we have estimated the median correlation yielded by the model for each connectivity value, as depicted in figure 6c for BB-networks and figure S3c for *α*-band networks, both for the RA and SOZ seeds. For BB-networks, the maximum overall correlations found were 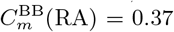 for *κ* = 36.90 for the RA seed; and 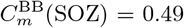 for *κ* = 19.68 for the SOZ seed. For *α*-band networks, the maximum overall correlations found are 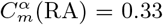 for *κ* = 19.68 for the RA seed; and 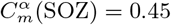 for *κ* = 19.68 for the SOZ seed. These values were smaller than the mean optimal results (*C*^BB^(RA) = 0.41, *C*^BB^(SOZ) = 0.51, *C*^*α*^(RA) = 0.38, *C*^*α*^(SOZ) = 0.47), but the decrease was less than 15% on average and not significant (*t*(9) = 1.95, *p* = 0.08 and *t*(9) = 1.90, respectively for the *α*-band and BB networks).

#### 3.2.4 Comparison with Fully Connected Networks

We also compared the correlation results with those obtained using a trivial fully connected network. Correlation results for this structure are shown in figures 7 and S4, together with the results obtained with individual broadband networks (averaged) and when using the average broad-band network. Although the fully connected network achieved a significant correlation for some patients (3 patients for the RA seed and 5 for the SOZ seed), the correlation was always lower than for the individually optimised model, except for one patient using the SOZ seed, and the average was significantly smaller (*C*^BB^(RA) − *C*^FCN^(RA) = 0.29, *p* = 1.4 · 10^*−*4^, *t*(9) = −6.27; *C*^BB^(SOZ) − *C*^FCN^(SOZ) = 0.22 *p* = 0.005, *t*(9) = 3.72; *C*^*α*^(RA) − *C*^FCN^(RA) = 0.27, *p* = 7 · 10^*−*4^, *t*(9) = 5.01; *C*^*α*^(RA) − *C*^FCN^(SOZ) = 0.18, *p* = 0.013, *t*(9) = 3.09).

### 3.3 Virtual Resection Analysis

We made use of the VR optimization method illustrated in figures 5 and S1 to find optimal virtual resections of increasing size *S*, for each patient. Results of the virtual resection analysis are shown in figure 8 for all patients. Propagation after the resection (as measured by *I*_*ℛ*_(*t*_0_)) decreased as *S* increased for all patients. However, the exact trend that was followed depended on the individual network structure and seed size. Patients 3, 5 − 7 showed a rapid decrease of *I*_*ℛ*_(*t*_0_)(*S*) for small *S*, whereas patient 9 showed a slower (parabolic) decrease. The remaining patients showed approximately linear decreases.

**Figure 8:**
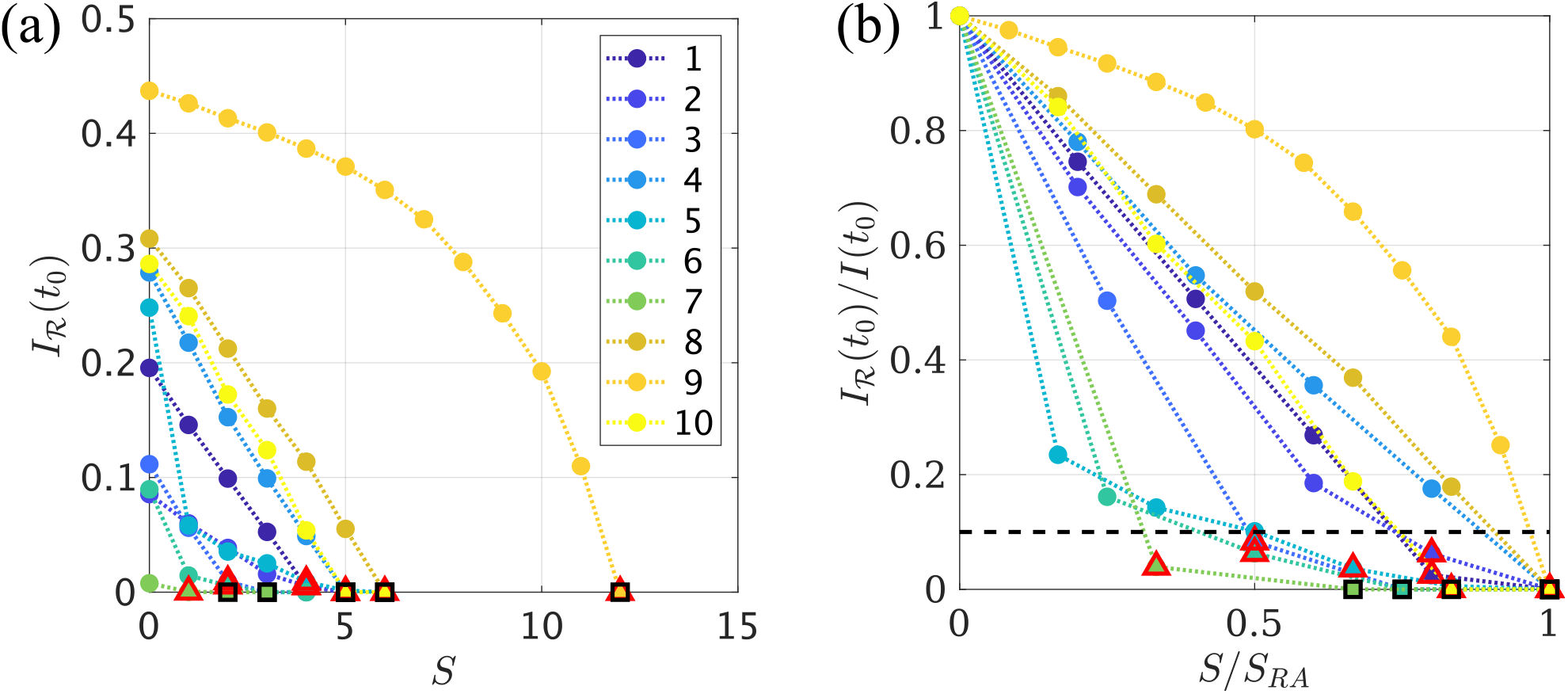
Optimal Virtual Resection. **(a)** Reduction in epidemic spreading for virtual resections of increasing size *S*, as quantified by the decrease in *I*_*ℛ*_(*t*_0_). Each curve corresponds to one patient, as indicated in the legend. Red triangles mark the resection that achieved a 90% decrease in propagation, *ℛ*_90_, and black squares the smallest resection that stopped seizure propagation, *ℛ*_100_. **(b)** In order to enable comparison of the VRs performance between patients, we depict the normalized decrease in propagation, *I*_*ℛ*_(*t*_0_)*/I*(*t*_0_) as a function of the normalized resection size, *S/S*_RA_. The black dashed line indicates a 90% decrease in propagation.

Complete stop of seizure propagation was found for the trivial resection of size *S* = *S*_RA_, which corresponds to complete removal of the seed, for all patients. However, in some cases the 100% resection ℛ_100_ (i.e. the smallest resection leading to a 100% decrease in seizure propagation) was smaller than resection area. This resection is indicated by black squares in figure 8. We found that ℛ_100_ whas smaller than the resection area for 4 patients (patients 4, 6, 7 and 10). Moreover, for 7*/*10 patients we were able to find a resection ℛ_90_, of smaller size than the actual resection, yet that achieved over 90% decrease in propagation, as indicated by red triangles in figure 8. The sizes of the ℛ_90_ and ℛ_100_ resections relative to the size of the resection area, i.e. 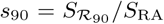 and 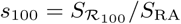, are shown in figure 9*a* for all patients. On average, *s*_90_ was 74% (range: 33 – 100%), whereas the *s*_100_ was 90% (range: 67 – 100%).

**Figure 9:**
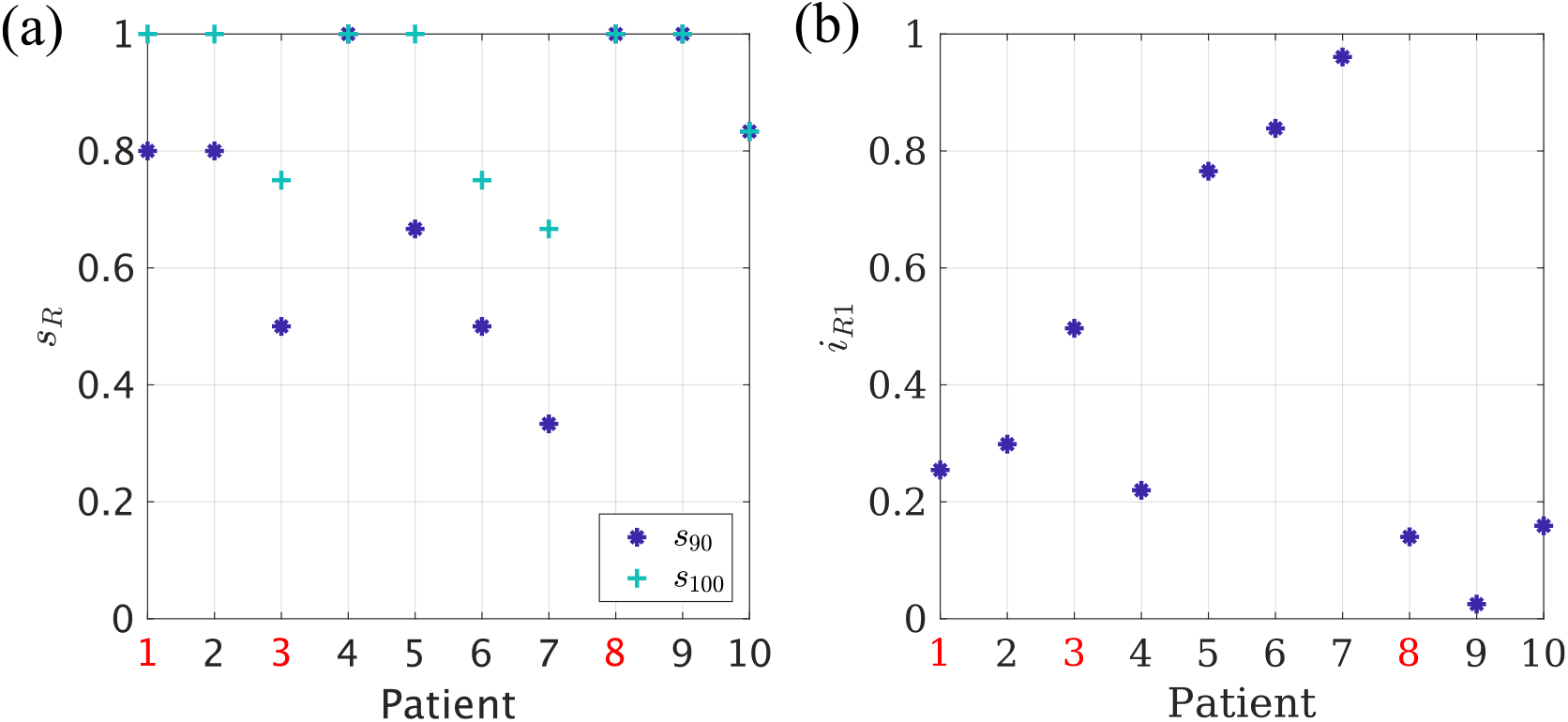
Analysis of optimal virtual resections. **(a)** Normalized size of the 90% (*s*_90_, dark blue asterisks) and 100% (*s*_100_, turquoise crosses) resections for each patient. **(b)** Normalized effect *i*_*ℛ*1_ = *I*_*ℛ*1_(*t*_0_)*/I*(*t*_0_) of the one node resection, for each patient. NSF patients are indicated by red labels in both panels.

The *I*_*ℛ*_(*S*) curves shown in figure 8 indicate that, for some patients, performing just a one-node resection, ℛ1, already had a large effect on (reducing) seizure propagation. This is explicitly shown by *i*_*ℛ*1_ = *I*_*ℛ*1_(*t*_0_)*/I*(*t*_0_) in figure 9*b*. The average effect of the 1-node resection was a 58% reduction in seizure propagation, although this number varied greatly among patients (range: 4–97%).

We analyzed the effect of the seed size and its connectivity on these results (see Supplementary Information, figure S5) by correlating *s*_90_, *s*_100_ and *i*_*R*1_ respectively with the number of links with nodes that were in the resection area, *E*_RA_ = *S*_RA_ * *κ*_RA_. We found that *s*_90_ and *s*_100_ correlated positively with *E*_RA_ (*r*(8) = 0.81, *p* = 0.005 and *r*(8) = 0.61, *p* = 0.07, for *s*_90_ and *s*_100_, respectively), although the correlation was only significant for *s*_90_. On the contrary, *i*_*ℛ*1_ correlated negatively with *E*_RA_ (*r*(8) = −0.71, *p* = 0.02). These results indicate that larger seed regions required a comparatively larger resection.

Finally, we analysed the location of the optimal resections found by the model (data not shown). We found that, for 1 patient, the optimal resection ℛ_90_ included nodes outside of the resection area and, similarly, ℛ_100_ was also found to include nodes outside of the resection area for another patient.

## 4 Discussion

We have defined a patient-specific seizure-propagation model based on the SI spreading dynamics. The model considers the patient-specific MEG functional connectivity matrix and makes use of clinical SEEG data to define stereotypical seizure propagation patterns for each patient. Seizure propagation was then modeled as an SI process propagating from a seed – which we initially took to be the patient’s resection area (RA) – to the rest of the network. Comparing the propagation patterns in the model to those observed clinically, we showed that this simple model reproduces the main aspects of the individual seizure propagation patterns, and that an alternative definition of the seed – based on the SEEG recordings – might provide a better reproduction of the observed propagation patterns. The main free parameter of the model – the network mean connectivity – was fitted to maximally reproduce the clinical seizure pattern, independently for each patient.

Using the model settings that optimally reproduced the clinically observed patterns, we then made use of the virtual resection technique to study alternative resections of smaller size or at different locations relative to the clinical resection area. The model suggested smaller virtual resections that were usually confined to the resection area, but in some cases included regions outside of the resected area.

### 4.1 Modeling Considerations

In this study we considered how individualized computer models, integrating patient-specific data from different modalities, can aid epilepsy surgery [26, 32–40, 42, 43, 78]. As opposed to previous studies which considered highly detailed, non-linear, stochastic models to simulate the activity of each brain region in detail [35, 37, 40, 43, 79–81], here we considered an abstract model of epidemic spreading, the SI model, as a proxy for seizure propagation dynamics (see figures 3 and 4). Epidemic models capture the basic mechanisms of processes that diffuse on networked systems, and have been used, for example, to study the propagation of pathological proteins on brain networks [52] and of ictal activity [41].

Moreover, epidemic models are supported by a well-grounded mathematical framework that can aid the exploration of the model. For instance, the fundamental role of hubs in seizure propagation is expected from a spreading perspective: hubs can act as super spreaders, being responsible for a disproportionate number of infections [54, 82], and their existence enhances epidemic spreading, both increasing the speed of propagation and decreasing the epidemic threshold [51]. On the contrary, a strong community structure can help control the epidemic, which may remain trapped in one community [83, 84]. This result also aligns with the clinical observation that often seizure propagation can be restricted to one or a few brain lobes [1], in the case of focal epilepsy. This is characterized by focal seizures that remain within some regions and only sometimes brake through the inhibitory “wall” and generalize. Interestingly, seizures originating in certain regions (such as the temporal lobes) are more likely to remain focal than others (such as frontal seizures). Similarly, other network characteristics such as temporal changes in connectivity [85, 86] or degree correlations can also alter behavior of spreading processes [87].

Epidemic models can thus help us studying seizure propagation processes. Of the large family of such models, we have selected the SI model as it captures the basic nature of epidemic spreading processes, including seizure propagation [51]. It only considers one mechanism: the propagation of an infectious process (or a seizure) from one region to another. Consequently, there is only one free parameter in the model – the probability that the infection is transmitted. This comes at the cost of not allowing for region deactivation: the model can only describe the initial steps of seizure propagation, when the activity starts to spread out. More detailed propagation models – such as the Susceptible-Infected-Susceptible (SIS) or Susceptible-Infected-Recovered (SIR) models – do include deactivation mechanisms, but in doing so extra parameters are introduced that would need to be fine tuned or assumed upon. Moreover, the early propagation-dominated phase of the SIR model is highly similar to the SI model, and this is the regime of interest here.

As the backbone for seizure propagation in the model, we used the broad-band AEC-MEG functional connectivity network as a proxy for the structural brain network (see figure 1). By not correcting for the effect of volume conduction/field spread, short-range connections are present in the network [70–73], yet it also captures long-range connections that might be difficult to capture with DTI-based tractography. Moreover, the functional network is a good indicator of how activity spreads on the network and, as we have shown, it suffices to reproduce the SEEG seizure propagation patterns when used in combination with the SI model.

### 4.2 Reproduction of seizure propagation patterns

In many patients, seizures follow stereotypical activation patterns. In this study we selected 10 patients who showed clear patterns on the SEEG recordings (see figure 2), and compared seizure propagation in the model with those clinically observed patterns, as depicted in figure 4. Despite its simplicity, we found that the model reproduces the main characteristics of the individual seizure propagation patterns in 9*/*10 patients when the resection area was considered as the seed (see figures 6a and 7a,b). Moreover, by using the possible seizure onset zone, as indicated by the SEEG recordings, as the seed for epidemic spreading, we showed that the model is sensitive to different definitions of the seed, and that alternative definitions can improve on the reproduction of the clinical patterns (see figures 6b and 7c,d). We also found that patient specific connectivity reproduces seizure propagation better than fully connected networks, and marginally (although not significantly) better than the average connectivity network (see figure 7). This result is in line with previous studies [36, 41] in which possible benefits of using patient-specific connectivity were suggested, but could not be corroborated by a significant difference in the model. Likely, larger data sets would be necessary to unravel how the models benefit from considering patient-specific connectivity.

The density of connections of the network was set for each patient to fit the SEEG seizure propagation pattern. Higher density levels imply a more extended or homogeneous propagation pattern, whereas smaller ones are associated with a more well-defined propagation. Then, the SI propagation rate *β* was adjusted accordingly for each patient for the subsequent virtual resection analysis.

The model parameters were fitted to the patient’s SEEG data, hence the current definition of the model relies on the use of SEEG recordings to infer the patient-specific seizure-propagation patterns and fit the model free parameters. However, these are not always part of standard clinical practice, as they are highly invasive for the patient and not always needed during pre-surgical planning. In order to show the feasibility of applying the model to patients without such recordings, we have shown in figure 6*c* that the average optimal model parameters can be used as an approximate solution. The propagation of seizures typically makes use of existing pathways, many of which are not patient-specific and can be recovered by the average model parameters. In the current setting, considering the overall best threshold for the connectivity matrix, instead of the individual one, led to less that a 15% decrease in correlation between the modeled and clinical seizure-propagation patterns. Within this configuration, the model is still personalized, as it is still fitted specifically for each patient via the patient’s MEG based connectivity matrix and seed for the SI propagation dynamics. Moreover, if a larger data-base is constructed, the patients could be grouped by epilepsy type and different optimal type-specific thresholds could be defined.

For seizure-free (SF) patients the resection area is, by definition, a better representation of the epileptogenic zone than for non-seizure-free ones (NSF). Thus, one might expect that modelled epidemics spreading from the resection area might also reproduce the clinically observed seizure propagation patterns better for SF patients than for NSF ones. In order to test this hypothesis, we compared the correlations between the modelled and the clinical propagation patterns for SF and NSF patients. We found no significant differences for any of the cases considered (i.e. using either the resection area or the SEEG-based SOZ as seed). The limited spatio-temporal resolution of the clinical propagation profile might be partially accountable for this result. The small group size (10 patients, only 3 of whom were NSF) prevents any further interpretations of this result. It is still worth noting, however, that in the current setting all patients had a big improvement in the frequency and severity of the seizures [2, 59], so the resection area provided a reasonable approximation for the EZ, even for the NSF patients.

### 4.3 Modelling Resections

The effect of different resections on seizure propagation can be studied with the model by implementing virtual resections (see figure 5). One can then search for optimal resections that minimize seizure propagation for a given resection size. This can be used to aid epilepsy surgery by either finding resections that are smaller than the standard clinical approach, but have the same or almost the same effect [41], to find alternative resections that avoid specific regions [40], such as eloquent cortex, or to propose alternative resections, including regions outside the hypothesized SOZ, that might lead to a better outcome for NSF patients.

The problem of optimization of virtual resections is highly computationally demanding. We found that a method that combines topological – using a surrogate structural measure [57, 58] – and dynamical properties can find optimal resections on the network. The use of this surrogate measure allows for a fast exploration of the space of possible resections, which is followed by a slower analysis using the SI dynamics to fine-tune the solution and measure the actual decrease in seizure propagation. In future studies exact results of the SI propagation on a network could also be implemented to avoid the need for random search methods [75, 88], taking advantage of the mathematical tractability of the SI model.

The effect of a resection in the model was measured as the decrease in propagation at a given time *t*_0_. That is, we measured the slowing down of seizure propagation due to the resection. In the model, a complete stop of propagation can only be achieved with – and is always achieved by – the complete disconnection of the seed from the rest of the network (this may not imply complete seed removal, in some cases removal of the nodes connecting the seed to the rest of the network might be enough and more efficient). This is because in the SI model activity always spreads to every connected region, eventually, regardless of any other network or model parameter. However, this model is only an approximation of actual seizure propagation, which is assumed to hold only for small times in which seizure dynamics are dominated by the activity propagation processes. After that, deactivation mechanisms kick in and the epileptiform activity eventually dies out. Within this paradigm, a sufficient decrease in seizure propagation at a given (early) time *t*_0_ would be enough to indicate an effective resection (that is likely to lead to seizure freedom); here this threshold was set to 90% of the original infection rate at time *t*_0_. Thus, we defined the optimal resection for each patient as the smallest resection leading to at least 90% decrease in seizure propagation. We found in the model that this optimal resection was smaller than the actual resection area for 7 out of 10 patients. Moreover, for four patients we found that it was possible to stop seizure propagation at the fixed time *t*_0_ with a resection smaller than the resection area. We found that cases with a larger or more densely connected epileptogenic region (i.e. a larger seed for the SI dynamics) required a larger portion of the seed to be removed to consistently reduce seizure propagation.

These findings highlight the need to devise patient-specific models to aid epilepsy surgery planning, so that optimized individualized resections with minimal side-effects can be found. In this study we allowed the search algorithm to consider nodes outside the resection area as targets for the virtual resections. We found that, for one patient, the optimal 90% resection included one node outside the RA, and similarly for another patient the 100% resection also included one node outside the resection area. Thus, individualized models could in some cases suggest alternative resections outside of the suspected epileptogenic zone that might be more beneficial than standard surgery.

The 90% threshold for the reduction in seizure propagation was set ad-hoc and was equal for all patients, which might not be realistic. Future studies could include a patient-specific estimation of the propagation threshold by analyzing the individual intracranial EEG recordings: often epileptiform activity appears in a confined region but does not propagate to the rest of the network. Information about the size of this region could be used to estimate the threshold for which a reduction in seizure propagation in the model is considered sufficient, that is, for which the modelled activity remains within this local region. Alternatively, de-activation mechanisms could be introduced in the dynamical model, such as in the SIR model [41]. By setting the model initially in the super-critical regime – in which seizures have a non-zero probability of propagating, the optimal resection would be the one that takes the model to the subcritical regime with the minimum resection size. However, the parameters of the model – namely the propagation and de-activation probabilities – would strongly affect this result: if the system is initially far into the super-critical regime, a larger resection will be necessary compared to when it is close to the critical transition. In fact, the SI model as it was used here corresponds to the highly-super critical case in which seizures always spread. Thus, in order to avoid more assumptions that would hinder the interpretation of the outcomes of such a model, the SIR parameters would need to be tuned with clinical data, using for instance high-resolution spatio-temporal seizure-propagation patterns [36, 44, 89].

#### 4.3.1 Alternative Resections for NSF patients

Another implication of the model definition is the fact that complete removal or isolation of the seed always leads to complete stop of seizure propagation, because seizures, in the model, only generate within the seed. Within the current formulation of the model, the seed is defined ad-hoc from the clinical data, either according to the resection area (RA seed) or the pre-surgical clinical information (SOZ seed). This implies that the resection model cannot distinguish situations where the selected seed is not a good representation of the EZ, such as is the case for NSF patients, and suggest a better resection. The first part of the model, i.e. the reproduction of seizure propagation patterns, could be of aid: the plausibility of different seeds can be judged from the maximum correlation that they yield between the modelled seizure patterns and the clinical SEEG patterns. Thus, optimal seed definitions that maximally reproduce the observed seizure propagation patterns could be suggested. This hypothesis implies that a higher correlation should be found for SF than for NSF patients when considering the resection area as the seed, which we were not able to validate in this study due to the small group size. Future studies should tackle this issue with larger patient groups and possibly more detailed spatio-temporal seizure propagation patterns, to increase the model resolution. Moreover, this would also validate whether the model can provide independent information prospectively – that is, prior to surgery – and suggest optimal seed definitions.

### 4.4 Strengths

The main strength of our approach is the simplicity of the model considered. Epidemic spreading models do not intent to capture the details of the underlying biological basis of seizure generation and propagation, only the stereotypical patterns of seizure propagation [78]. The simplicity of the model not only allows for faster calculations and fewer free parameters, but it also comes with a large body of theoretical and computational studies that can be used to interpret the results and design the study [51].

We integrated data from different modalities that are commonly measured in clinical practice: MEG and SEEG recordings, and the location of the resection area. The use of MEG networks as a proxy for structural networks avoids the computation of structural DTI networks, which are not part of standard clinical care and also time-consuming, limiting the flexibility with which the choice of atlas can be changed. Using MEG functional networks to define the backbone for the dynamical model allows for more versatility, as well as the ability to use our approach in patients for whom DTI data are not available.

The model was fitted with patient-specific data and optimized independently for each patient. The varying results for different patients, both for the reproduction of seizure propagation patterns and the analysis of alternative resections, highlight the need for using personalized models of seizure propagation [36, 41, 89].

Moreover, the model could be easily extended to include more clinical presurgical information, such as the existence of MRI or MEG abnormalities. Similarly, the model could be used prospectively by using alternative definitions of the seed, that do not depend on the resection area, as we have already shown here by using seeds based on the SOZ as determined from SEEG (see figures 6 and 7).

### 4.5 Limitations

The main limitations of the current study are the limited number of patients considered and the low-resolution of the clinical seizure propagation patterns. The small cohort prevents further validation of the model to distinguish between SF and NSF patients. Meanwhile, the low resolution implies that few parameters of the dynamical model can be fit to the data.

Another important limitation is that the seed of seizure propagation was assumed from the data – being either the resection area or the SOZ as estimated from the SEEG recordings. This, in conjunction with the fact that complete removal of the seed always leads to a stop of seizure propagation, implies that the model cannot suggest better resections for NSF patients, and it also limits its prospective use. To be of more clinical use, the model should be able to suggest the seed of seizure propagation. This could be done by finding the set of nodes that maximally reproduces the clinical seizure propagation patterns. However, in order to do this realistically and in a systematic manner, more detailed spatio-temporal patterns of seizure propagation are needed for each patient. These could be obtained from the SEEG recordings directly [36, 44, 89].

Another limitation is the nature of the SI model: it reproduces adequately the initial steps of seizure propagation, but the lack of a de-activation mechanism means that it cannot fit the whole seizure. Including a mechanism for de-activation would circumvent this issue, provided that the extra parameters can be adequately fitted.

The use of SEEG data to fit the model can be another limitation for its clinical use, as SEEG recordings are highly invasive and not always part of the presurgical evaluation. However, we have shown that the model parameters can be extrapolated from the overall best fit (see figure 6c) and used for patients without SEEG. In the current setting, this led to a less than 15% decrease in the reproducibility of the seizure patterns, as measured by the correlation between the SI and SEEG propagation patterns. Information from other modalities could also potentially be included, such as epileptiform abnormalities found in MEG imaging. These can be used to set the probability for a region being a seed region.

An inherent limitation of all studies analyzing the functional effect of different resections is modeling the resection itself. Here we have employed the commonly used *virtual resection* technique, such that the weights of all resected links are set to 0 [37, 79, 90, 91]. This does not account for the generalized effect that a local resection can have on the network [92]. It does not consider any plasticity mechanisms either [93, 94], which are known to occur following a lesion in the brain [11, 95] – and in particular following a resection [96–99]. These appear as a consequence of the network disruption, and can have widespread effects. They will play a significant role in the cognitive functioning following the resection, and can also affect the long-term outcome of the surgery. These effects should be included in future studies for a more comprehensive modelling of epilepsy surgery.

A final limitation of the study, and of similar studies using the virtual resection technique, is the difficulty of the validation of the results, as the different resections cannot be tested clinically. Typically, virtual resection models are validated by comparing the overlap between the suspected EZ as generated by the model with the resection area for both SF and NSF patients, where a valid model should provide a good match for SF patients and a poor match for NSF patients [32, 35, 37, 38, 42]. Alternatively, the propagation pathways simulated by the model are compared with those recorded with SEEG [36, 89]. We have undertaken the later approach in this work to tune the model parameters and for validation, and the first approach was used for validation, although the small group sizes do not allow us to draw strong conclusions in this proof-of-principle study. Moreover, using surgical outcome to validate the model is only a first step, as the ultimate goal is to improve surgical outcome, i.e. to perform the analysis proposed in this work before surgery has taken place.

### 4.6 Outlook

In recent years there have been increasing efforts to develop individualized computer models to study brain disorders. In particular, in the case of epilepsy surgery, it is expected that such models might help improve surgery outcome and decrease the cognitive side-effects associated with epilepsy surgery, by proposing targeted, individualized resections for each patient. Currently, the greatest challenge remains in the validation of the models, as the ground truth is inherently missing and the actual effect of a resection can only be known several months – or years – after the surgery has taken place. Thus, extensive retrospective validation of the models is necessary before prospective (or even pseudo-prospective) studies can take place. Here, the seizure propagation model ought to be validated in future studies by increasing the number of included patients, and the resolution of the SEEG seizure propagation pattern should be increased in order to increase the sensitivity to the model parameters. Then, if a better relation between the model and the clinical data can be found for SF than for NSF patients, as we have hypothesized, the model could be used to find the seed regions that maximally reproduce the seizure patterns. Similarly, future studies could explicitly include forbidden areas that cannot be removed during the surgery, such as the eloquent cortex, or avoid surgeries that are not possible in clinical practice.

## 5 Conclusions

Patient-specific epidemic models can capture the fundamental aspects of seizure propagation as observed clinically with invasive SEEG recordings. The models, optimized specifically for each patient, can then be used to test the effect that different resection strategies may have on seizure propagation *in silico*. Our results highlight the need for individualized computer models to aid epilepsy surgery planning by defining smaller targeted resections with potentially fewer side-effects and better outcome than standard surgery.

## Data Availability

The data used for this manuscript are not publicly available because the patients did not consent for the sharing of their clinically obtained data. Requests to access to the datasets should be directed to the corresponding author.
All user-developed codes are available from the corresponding author upon reasonable request.

## 6 Acknowledgements

Ana P. Millán and Ida A. Nissen were supported by ZonMw and the Dutch Epilepsy Foundation, project number 95105006. The funding sources had no role in study design, data collection and analysis, interpretation of results, decision to publish, or preparation of the manuscript.

## 8 Author Contributions

A.P.M., E.C.W.S., C.J.S., I.A.N, A.H. conceptualized the study, E.C.W.S., C.J.S., I.A.N, S.I., J.C.B., P.V.M., A.H. participated in the funding acquisition, A.P.M, E.C.W.S., C.J.S, A.H. devised the Methodology, A.P.M. performed the formal analysis, A.P.M, I.A.N, A.H. devised the software and visualization E.C.W.S., C.J.S., P.V.M., A.H. participated in the supervision, E.C.W.S., S.I., J.C.B. provided resources, A.P.M. wrote the original draft and all authors participated in writing review and editing.

