## Supplementary Information for "Epidemic models characterize seizure propagation and the effects of epilepsy surgery in individualized brain networks based on MEG and invasive EEG recordings"

#### 1 Optimization of virtual resections

For a given resection size $S_{\mathcal{R}}$ (number of resected nodes), the optimal virtual resection $\mathcal{R}^*(S_{\mathcal{R}})$ is the one that creates the largest reduction in seizure propagation, $\Delta^*_{\mathcal{R}}(S_{\mathcal{R}})$. Because characterizing all possible virtual resections is a combinatorial problem that soon looses tractability, alternative approaches are necessary to find the optimal solution. Therefore, in order to alleviate the exploration of the parameter space, we defined a random search method that combined structural and dynamical information, and made use of a simulated annealing algorithm. The method considers a surrogate topological measure for the SI dynamics, which is used to fasten up the exploration of the space of virtual resections, and the solutions are subsequently refined using the actual SI dynamics. The optimization method is described in detail below, and a schematic view is given in figure $S1$.

##### 1.1 Surrogate model

In the SI model, the probability that the infection has reached a certain node $i$ at a given time $t$ decays in an approximate linear manner with $d_i^{\text{seed}}$[1], where $d_i^{\text{seed}}$is the average distance from the considered node to the seed (see figure $S1$). That is,

$$d_i^{\text{seed}} = S_{\text{seed}}^{-1} \sum_{j \in \text{seed}} d_{ij}, \tag{S1}$$

where $d_{ij}$ is the topological distance between the pair of nodes $(i, j)$ and $S_{\text{seed}}$ is the number of nodes in the seed region. $d_{ij}$ is defined as

$$d_{ij} = \sum_{k \in \ell_{ij}} \ln(\frac{w_k}{w_0}), \tag{S2}$$

where the sum is over the edges in the shortest path between nodes $i$ and $j$, $w_k$ is the weight of edge $k$ and $w_0$ is the minimum non-zero weight, introduced for normalization purposes [1]. The average distance to the seed, $d^{\text{seed}} = (N - S_{\text{seed}})^{-1} \sum_i d_i^{\text{seed}}$, where the sum is over nodes that do not belong to the seed and $N = 246$ is the total number of nodes in the network (i.e. the number of ROIs), can be used as a topological proxy for the propagation of the SI dynamics. The topological measure that quantifies the effect of a given virtual resection is thus the increase in the average distance to the seed:

$$\Delta\mathcal{R}_{\mathcal{T}} = d_{\mathcal{R}}^{\text{seed}} - d^{\text{seed}}. \tag{S3}$$

#### 1.2 Optimization method

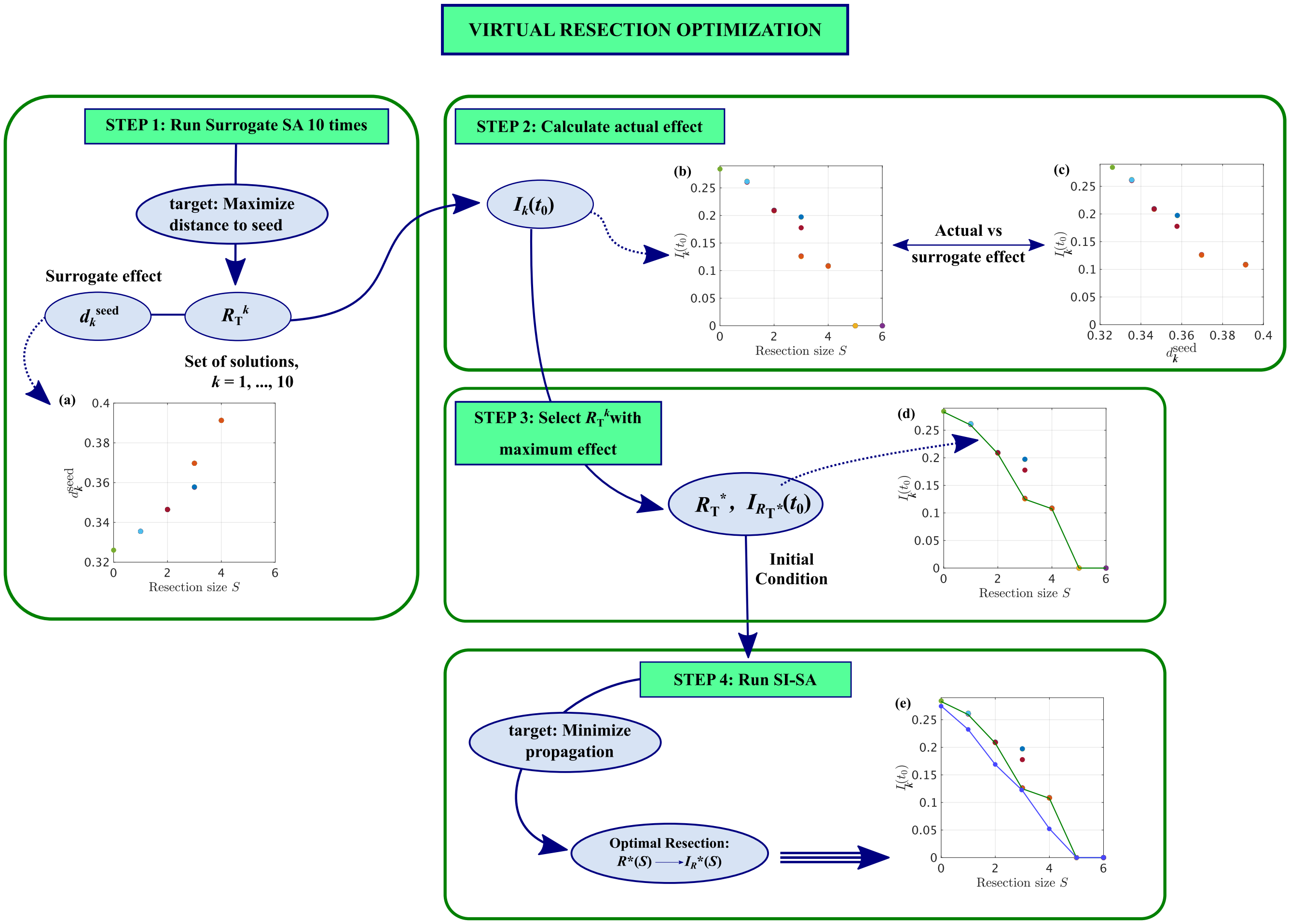


Figure S1: **Schematic representation of the Virtual Resection Optimization Method.** We used a four step optimization approach to find the virtual resection that, for a given resection size, minimized seizure propagation. Firstly (step 1), we used the SA algorithm to find resections that maximized the surrogate measure $\Delta\mathcal{R}_{\mathcal{T}}$ (eq. $S3$). The optimization algorithm was iterated 10 times, yielding a set of 10 possible virtual resections, $\mathcal{R}^k_{\mathcal{T}}(S)$, $k = 1, ..., 10$, for each resection size. This is depicted in panel **(a)**, where we show $d_k^{\text{seed}}$ as found by the SA algorithm for VRs of different size. In most cases all solutions had the same $d_k^{\text{seed}}$ and overlap in the figure. For this case, the seed became completely disconnected for $S = 5$, which is 1 node less than the size of the RA. Next (step 2), we calculated the actual effect of each resection $\mathcal{R}^k_{\mathcal{T}}(S)$, $I_k(t_0)$, as depicted in panel **(b)**, where we show $I_k(t_0)$ for those same resections (error bars indicating the standard deviation among 10 iterations of $10^4$ runs of the SI dynamics overlapped with data points and are not visible). Panel **(c)** illustrates the relationship between the surrogate and actual effects of each resection, i.e. between the topological measure $d_k^{\text{seed}}$ and dynamical one $I_k(t_0)$, for each VR. $I(t_0)$ decayed roughly proportionally with $d^{\text{seed}}$until it saturated and equaled 0 for complete seed isolation (in which case $d^{\text{seed}}$diverged, not shown here). In step 3, the resection leading to the maximum actual effect for each resection size, $\mathcal{R}^*_{\mathcal{T}}(S)$, was selected as shown in panel **(d)**. Finally, for each resection size separately, this resection was used as the initial condition for the SA algorithm to minimize seizure propagation, as measured by $I_{\mathcal{R}}(t_0)$ (panel **(e)**). This led to the final optimal resection, $\mathcal{R}^*(S)$, characterized by the propagation $I^*_{\mathcal{R}}(S)$, for each resection size. In this case, we found that the propagation was stopped for $S = 5$.

In order to find optimal resection strategies, and considering the strong non-linearities present in the model, we have made use of a probabilistic optimization algorithm, *Simulated Annealing* (SA) [2], to optimize the effect of virtual resections, for a given number of resected nodes. In order to do so, we employed a four-step method to find, for each resection size $S$ up the size of the resection area, $S_{\mathrm{RA}}$, the resection leading to the maximum decrease in seizure propagation (see figure $S1$):

1. Firstly, we considered the surrogate model, and used the SA algorithm to maximize $\Delta\mathcal{R}_{\mathcal{T}}$. The SA algorithm is probabilistic, and was therefore run 10 times, yielding a set of surrogate solutions $\mathcal{R}_{\mathcal{T}}^{k}$, with $k = 1, \ ..., \ 10$, each characterized by its topological effect $\Delta\mathcal{R}_{\mathcal{T}}^{k}$, as shown in figure $S1$a. Figure $S2$ shows an exemplary case of the SA optimization search.

2. The topological metric $\Delta\mathcal{R}_{\mathcal{T}}$ is only an approximation of the actual effect each resection $\mathcal{R}$ has on the propagation dynamics. Consequently, for each surrogate solution $\mathcal{R}_{\mathcal{T}}^{k}$, its actual effect was then determined by estimating $I_{\mathcal{R}_{\mathcal{T}}}^{k}(t_0)$ (figure $S1$b), i.e. seizure propagation on the network once the virtual resection has taken place.

3. The optimal topological resection of size $S$, $\mathcal{R}_{\mathcal{T}}^{*}(S)$, was defined as the one leading to the maximum decrease in epidemic spreading (figure $S1$c).

4. Finally, we made use of the SA algorithm again: the optimal topological resection $\mathcal{R}_{\mathcal{T}}^{*}(S)$ was used as the initial condition for the search. This time, the algorithm was used to minimize $I_{\mathcal{R}}(t_0)$ directly (see figure $S1$d). This yielded the final optimal resection $\mathcal{R}^{*}(S)$, with propagation $I_{\mathcal{R}}^{*}(S)$.

This method was iterated from $S = 1$ up to the size of the resection area, $S = S_{\mathrm{RA}}$. Overall, this method allows for a faster exploration of the parameter space (using the surrogate model), and a posterior refinement of the solution using the seizure propagation model.

##### 1.3 SA Algorithm

We used a MATLAB implementation of the Simulated Annealing (SA) algorithm (J. Vandekerckhove, general simulated annealing algorithm, version 1.0.0.0, MATLAB Central File Exchange), adapted for our model (see figure $S2$). Starting out with a given selection of nodes for the virtual resection, at each step of the SA algorithm this set is updated by changing $n$ of the nodes. $n$ follows a power-law distribution to allow for non-linear exploration of the node-space. This SA algorithm minimizes a cost function, which was defined as the inverse of $d_{\mathcal{R}}^{\mathrm{seed}}$ for the surrogate case (step 1 in figure $S1$), and as the fraction of infected nodes at $t_0$, $I_{\mathcal{R}}(t_0)$ for the final step (step 4 in figure $S1$).

The other parameters of the algorithm were set to default for step 1: initial temperature = 1, stopping temperature = $10^{-8}$, annealing function $f(T) = 0.8T$, maximum number of consecutive rejections = $10^3$, maximum number of tries within one temperature = 300, maximum number of successes within one temperature = 20. The algorithm was run 10 times with different (random) initial configurations, and the one with the largest effect was selected. For step 4, some parameters were changed to allow for a faster approach to the final solution: stopping temperature = $10^{-6}$, annealing function $f(T) = 0.6T$. In this case the algorithm was only run once for each resection size, using the optimal solution from the surrogate SA method as the seed.

Due to the random nature of the SI dynamics, different realizations with the same seed can lead to different outcomes. The SA algorithm could thus artificially minimize $I(t_0)$ by selecting realizations that, by chance, have a smaller propagation at $t_0$. Averaging over a large number of realizations ($10^4$ in the current case) minimizes this problem, but it can still occur. To avoid this artificial minimization, after accepting a new virtual resection $\mathcal{R}$ the propagation $I_{\mathcal{R}}(t_0)$ was calculated again with a new simulation of the SI dynamics. In this manner the realizations that were used to select a resection were not carried over in subsequent iterations. This step was not necessary in case of the surrogate model since the calculation of $d_{\mathcal{R}}^{\mathrm{seed}}$ is exact.

#### 2 Reproduction of seizure propagation patterns: Results.

##### 2.1 Optimal correlation for BB networks.

In table $S1$ we report the optimal correlation found for each patient using the patient-specific BB-connectivity network, and the corresponding value of the mean connectivity.

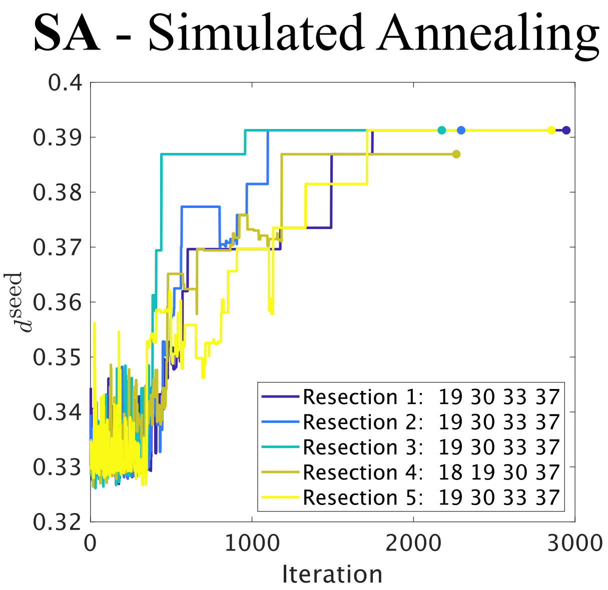


Figure S2: **Surrogate Simulated Annealing.** Here we illustrate the Simulated Annealing (SA) method, for an exemplary VR of size $S = 4$, for patient 10. The SA algorithm performs a random search, starting from a random virtual resection, so that at each step a new resection is calculated by switching some of the selected nodes. The new resection is then accepted with a probability that depends on the difference of effect between the resections, so that resections that increase $d^{\text{seed}}$have a higher probability of being accepted. This process is iterated until a stable solution is found. Due to its probabilistic nature, different realizations of the algorithm can lead to different outcomes, of which the optimal one is selected. In this case, iterations 1, 2, 3, 5 reached the same resection (target nodes 19, 30, 33, 17) that maximally increased $d^{\text{seed}}$.

| | RA Seed | | SOZ Seed | | |
|---|---|---|---|---|---|
| Patient | Correlation | $\kappa$ | Correlation | $\kappa$ | Outcome |
| P1 | 0.48* | 36.90 | 0.65* | 14.75 | NSF |
| P2 | 0.62* | 24.60 | 0.52* | 19.68 | SF |
| P3 | 0.34* | 14.75 | 0.30* | 14.75 | NSF |
| P4 | 0.41* | 49.20 | 0.53* | 36.91 | SF |
| P5 | 0.40* | 9.84 | 0.49* | 9.84 | SF |
| P6 | 0.36* | 4.92 | 0.53* | 9.84 | SF |
| P7 | 0.14 | 4.92 | 0.62* | 14.75 | SF |
| P8 | 0.43* | 49.20 | 0.48* | 24.60 | NSF |
| P9 | 0.53* | 14.75 | 0.53* | 14.75 | SF |
| P10 | 0.43* | 9.85 | 0.48* | 9.84 | SF |
| Mean (all) | 0.41 | 21.89 | 0.51 | 16.97 | |
| Mean (SF) | 0.41 | 16.87 | 0.53 | 16.52 | |
| Mean (NSF) | 0.42 | 33.62 | 0.48 | 18.03 | |

Table S1: **Model optimization.** For each patient the maximum correlation between the clinical and model seizures patterns, and the corresponding optimal network connectivity, for the broad-band (BB) networks, is given, both for the RA and SOZ seeds. The seizure outcome is also indicated. The final three rows indicate the average correlation and density obtained for the whole population (all), seizure free patients (SF) and not seizure free patients (NSF). * Indicates a significant correlation ($p < 0.05$ ).

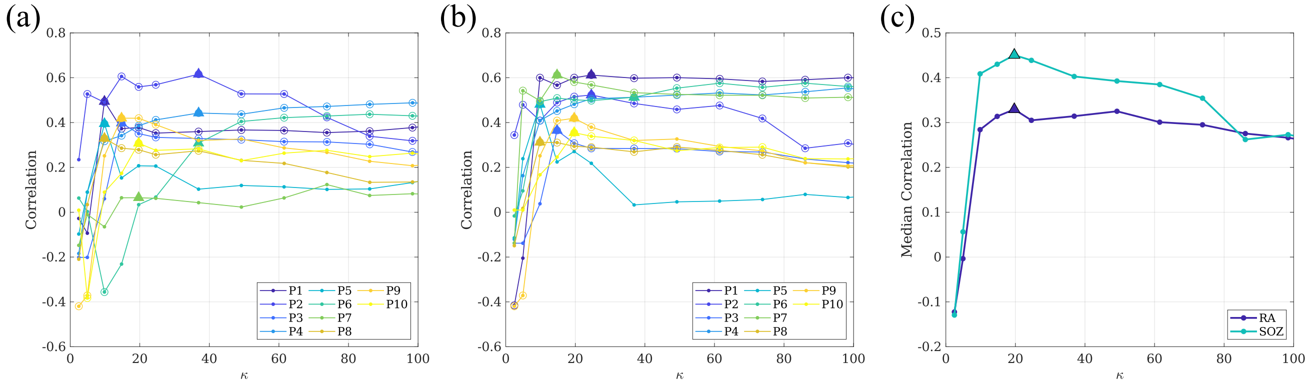


Figure S3: **Reproduction of seizure propagation.** Correlation between the modelled and clinically observed seizure propagation pattern as a function of network density, for each patient, using the RA (panel **a**) and SOZ seeds (panel **b**), for the alpha-band networks. Panel (**c)**) indicates the median curves for each case, as indicated by the legend. Circles in panel a and b denote significant correlations ($p < 0.05$), and the optimal correlation for each case is marked with a triangle.

#### 2.2 Results for $\alpha-$band networks.

We report here on the results of the seizure propagation model fit and validation analysis for the $\alpha$-band networks. As described in the methods section of the main text, we measured the correlation $C$ between the SI and SEEG propagation patterns for different values of the mean network connectivity $\kappa$. The resulting $C(\kappa)$ curves are shown in figure $S3$, in panels a and b for the RA and SOZ seeds, respectively. As it was the case for the BB networks, we found a bimodal dependence of $C(\kappa)$ for most patients, so that there is a maximum for low density values and another one for large densities. As before, we restricted our considerations to the first maximum, which leads to networks with only the fundamental connections that are needed to reproduce seizure propagation. Then, we found the connectivity value $\kappa_{\text{max}}$ that led to the maximum correlation, independently for each patient. We found that the model significantly reproduced the seizure propagation patterns for 9 and 10 out of 10 patients for the RA and SOZ seeds, respectively. The maximum correlation $C_{\text{max}}$ found for each patient is indicated in figure $S3$ with a triangle, and the corresponding values are reported in table $S2$ and figure $S4$.

We also report in figure $S4$ the optimal correlation found when using the average $\alpha$-band network and the fully connected network. The average values for each group are reported on the main text. Finally, in figure $S3$c we show the average model as given by the median correlation of $C(\kappa)$ for all patients, for both the RA and SOZ seeds.

### 3 Analysis of VR results

Figure $S5$ reports the dependence of the normalized size of the 90% and 100% (panel a) and of the normalized effect of the 1-node resection (panel b) on the number of links of the resection area, $E_{\text{RA}}$. $E_{\text{RA}}$ combines the effect of the size and connectivity of the resection area. We found that $s_{90}$ and $s_{100}$ correlated positively and significantly with $E_{\text{RA}}$ ($r(8) = 0.81$, $p = 0.005$ and $r(8) = 0.61$, $p = 0.07$, respectively for $s_{90}$ and $s_{100}$) although the correlation was only significant for $s_{90}$. We analyzed whether this effect was led by the outlier case with largest $E_{\text{RA}}$ by removing it from the correlation analysis. The results ($r(7) = 0.85$, $p = 0.004$ and $r(7) = 0.64$, $p = 0.06$, respectively for $s_{90}$ and $s_{100}$) indicate that this was not the case case. $i_{R1}$ correlated negatively and significantly with $E_{\text{RA}}$ ($r(8) = -0.71$, $p = 0.02$), and the effect was also not led by the outlier case ($r(7) = -0.65$, $p = 0.05$). Thus, larger or more densely connected epileptogenic regions would require larger minimal resections, as expected.

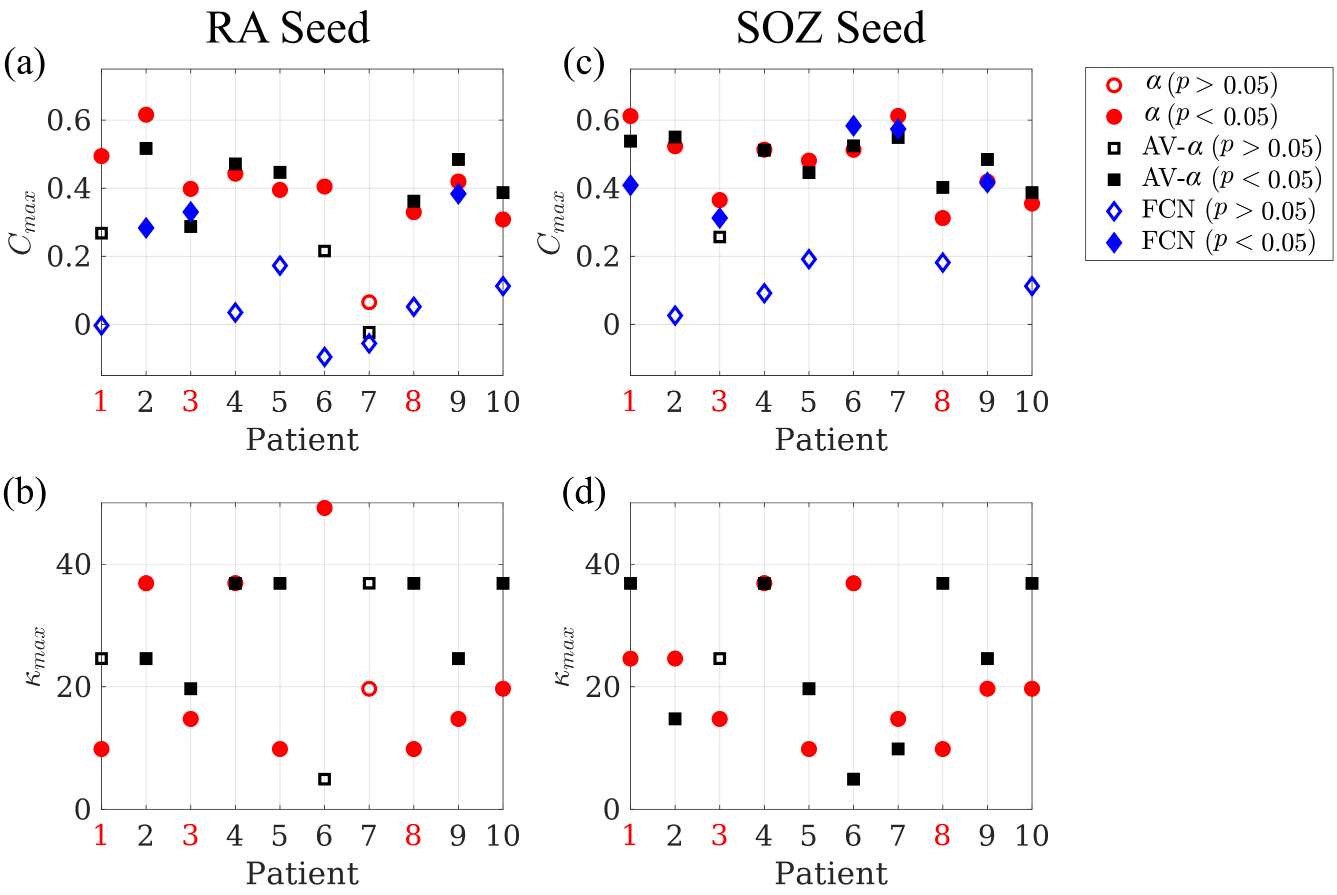


Figure S4: **Reproduction of seizure propagation patterns.** Panels **(a)** and **(c)** show the average maximum correlation $C_{max}$ achieved by the individual $\alpha$-band networks ($\alpha$, red circles), and the average AB one (AV-BB, black squares), and the correlation found for the fully connected network (FCN, blue diamonds), respectively for the RA and SOZ seeds. Panels **(b)** and **(d)** show the corresponding $\kappa_{\max}$ for the individual (AB, red circles) and average (AV-AB, black squares) networks. Significant correlations ($p < 0.05$) are indicated by a filled marker. NFS patients are indicated by red labels.

| | RA Seed | | SOZ Seed | | |
|---|---|---|---|---|---|
| Patient | Correlation | $\kappa$ | Correlation | $\kappa$ | Outcome |
| P1 | 0.49* | 9.84 | 0.61* | 24.60 | NSF |
| P2 | 0.62* | 36.90 | 0.52* | 24.60 | SF |
| P3 | 0.40* | 14.75 | 0.36* | 14.75 | NSF |
| P4 | 0.44* | 36.90 | 0.51* | 36.90 | SF |
| P5 | 0.39* | 9.84 | 0.48* | 9.84 | SF |
| P6 | 0.31* | 36.90 | 0.51* | 36.90 | SF |
| P7 | 0.06 | 19.68 | 0.61* | 14.75 | SF |
| P8 | 0.33* | 9.84 | 0.31* | 9.84 | NSF |
| P9 | 0.42* | 14.75 | 0.42* | 19.68 | SF |
| P10 | 0.31* | 19.68 | 0.35* | 19.68 | SF |
| Mean (all) | 0.38 | 20.90 | 0.47 | 21.15 | |
| Mean (SF) | 0.36 | 24.90 | 0.49 | 23.19 | |
| Mean (NSF) | 0.41 | 11.50 | 0.43 | 16.39 | |

Table S2: **Model optimization.** For each patient, the maximum correlation between the clinical and modelled seizures patterns, and the corresponding optimal network connectivity, for the $\alpha-$band networks, is given for the RA and SOZ seeds. The seizure outcome is also indicated. The final three rows indicate the average correlation and density obtained for the whole population (all), seizure free patients (SF) and not seizure free patients (NSF). * Indicates a significant correlation ($p < 0.05$ ).

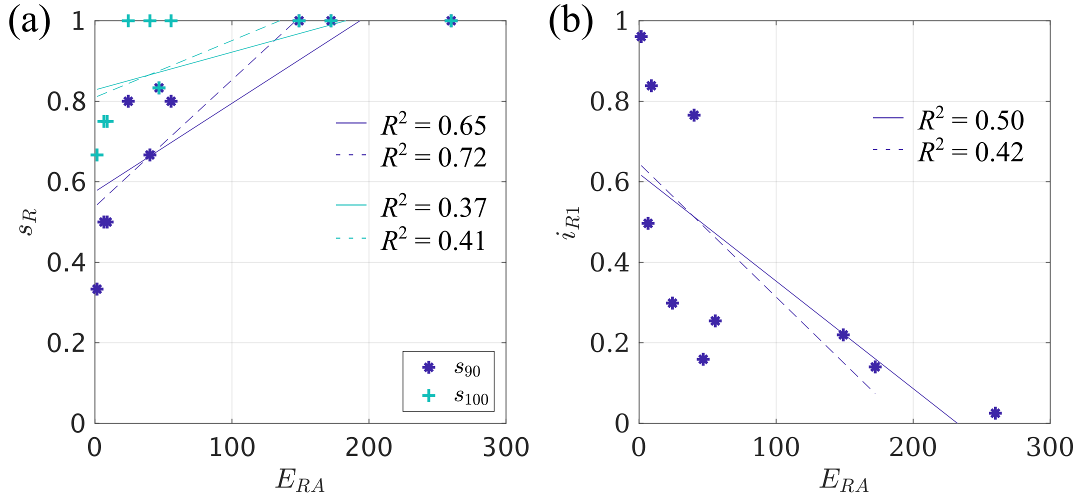


Figure S5: **Analysis of VR results. (a)** Dependence of the normalized size of the 90% and 100% resections (respectively $s_{90}$ and $s_{100}$) on the number of links of the resection area, $E_{\mathrm{RA}}$. **(b)** Dependence of the normalized effect of the 1-node resection on $E_{\mathrm{RA}}$. Solid lines indicate the fits to all data points, and dashed lines the fits without the outlier case, for both panels.

[2] S. Kirkpatrick, C. D. Gelatt, and M. P. Vecchi. "Optimization by simulated annealing". *Science* 220.4598 (1983).
